# Gold Nanourchins Improve Virus Targeting and Plasmonic Coupling for Virus Diagnosis on a Smartphone Platform

**DOI:** 10.1101/2022.08.25.22279227

**Authors:** Yaning Liu, Haihang Ye, Abdullah Bayram, Tingting Zhang, Qi Cai, Chen Xie, HoangDinh Huynh, Saquib Ahmed M. A. Peerzade, Jeffrey S. Kahn, Zhenpeng Qin

## Abstract

Point-of-care detection of pathogens is critical to monitor and combat viral infections. Here, we demonstrate a plasmonic coupling assay (PCA) using gold nanourchins (AuNUs) as labels for the colorimetric quantification of viruses. The antibody functionalized AuNUs allow for rapid and highly specific identification of viruses and provide strong color change for sensitive detection. Using respiratory syncytial virus (RSV) as a target, we demonstrate that the AuNU-based PCA achieves a detection limit of 1,402 PFU/mL (equivalent to 17 copies/μL) that is 3.1- and 5.7-times lower than the rod- and sphere-based counterparts, respectively. The improved detection sensitivity arises from the higher virus binding capability and stronger plasmonic coupling at long distances (∼10 nm) by AuNU probes. The detection can be performed with a portable smartphone-based spectrometer and is validated by testing RSV-spiked nasal swab clinical samples. Our study reports a rapid and sensitive approach for intact virus detection and provides a potential toolkit at the point of care.

**Graphical abstract:** 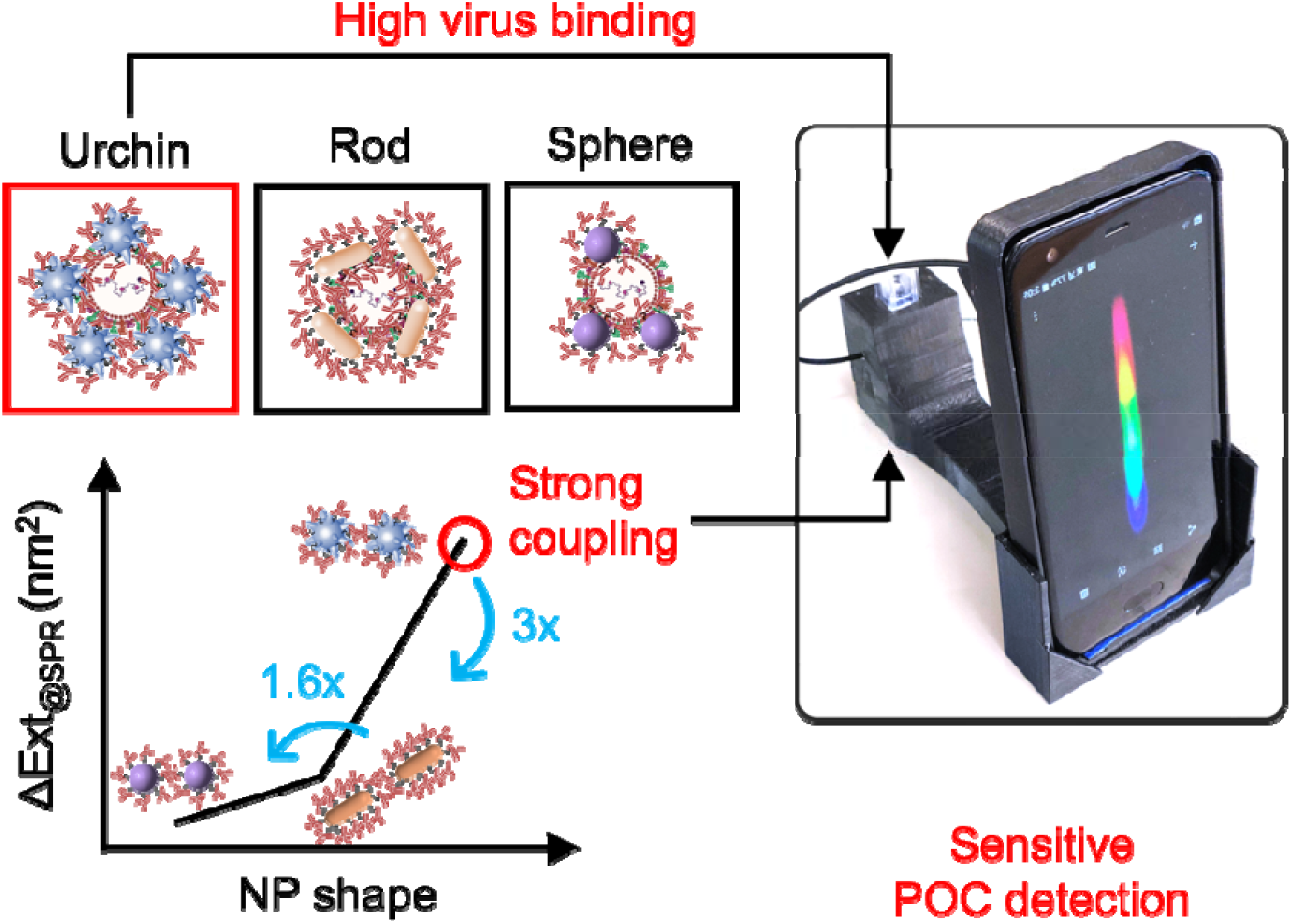

## 1. Introduction

Respiratory infectious diseases have significantly burdened millions of people each year, especially in the current COVID-19 pandemic.^1^ Among them, respiratory syncytial virus (RSV) is one of the leading causes of lung diseases among infants and young children under 5 years of age.^2^ Currently available RSV detection methods include laboratory tests^3^ (e.g., polymerase chain reaction and viral culture assays) and rapid antigen tests (e.g., lateral flow assay^4^ and direct fluorescent antibody testing^5^). While sensitive and specific, laboratory tests have a long turnaround time and are inaccessible in resource-limited settings.^6^ In contrast, rapid antigen tests suffer from low sensitivity and are not recommended for adults and elder children.^7^ Therefore, there is an unmet and urgent need for simple and sensitive detection of RSV.

Homogeneous assays are simple and rapid detection methods without the need for immobilization, separation, and washing, and thus are promising for point-of-care (POC) diagnosis.^8^ Plasmonic nanomaterials, such as gold and silver nanoparticles (Au and AgNPs), have been employed for homogeneous assays (termed as plasmonic coupling assays, PCAs) due to their unique localized surface plasmon resonance (LSPR) properties.^9, 10^ Taking the AuNPs as an example, they show vivid colors in the visible region (400-800 nm) and have over 3 orders of magnitude higher extinction coefficients than organic chromophores.^11^ Upon binding to the target and in close proximity to each other, they display color changes. Additional merits include good stability and easy surface modification for bio-molecule conjugation (e.g., via thiol bond).^12^ Since the first report by Mirkin et al.,^13^ various AuNPs-based PCAs have been developed with advantages of simplicity, convenience, low cost, and visual detection. However, PCAs have so far suffered from low detection sensitivity for intact pathogen detection in clinical applications, making them inadequate as a standalone diagnostic tool. Previous efforts focused on the additional signal amplification strategies based on field enhancement such as surface-enhanced Raman spectroscopy (SERS) and sample enrichment such as multiple washing or purification steps (see comparison in **Table S1**). Although sensitive, they added additional complexity for the assay including the need of laboratory infrastructures, and hence have limited applications. Considering the optical properties of plasmonic NPs are highly dependent on their geometry (e.g., size and shape),^14^ engineering plasmonic AuNP probes offers a potential opportunity to further improve the PCA performance.

In this study, we systematically investigated the effect of particle shape on the AuNP-based PCA with the goal to substantially improve the detection sensitivity. We evaluated the PCA performance using AuNPs with spherical, rod, and urchin-like shapes for the colorimetric detection of RSV. We demonstrated that Au nanourchins achieve a limit of detection (LOD) of 1,402 PFU/mL (equivalent to 17 copies/μL), which is significantly lower than that of nanorod and nanospheres. By electron microscopy imaging of the nanoparticle-virus binding, we revealed that rod and urchin nanoparticles bind to the virus particle more effectively than spherical nanoparticles. Furthermore, modeling of the plasmonic interactions shows that coupled Au nanourchins show the highest plasmonic coupling (i.e., the largest change in the extinction spectrum) at long distances (∼10 nm) that represent the distances for immunorecognition. We further demonstrated that the detection can be performed on a smartphone-based spectrometer and the single-step detection works on clinical specimens spiked with RSV. With improved sensitivity and portability and a simple one-step assay, our point-of-care diagnostic platform has great potential to facilitate virus detection at its early presentation and combat viral infections (**Scheme 1**).

**Scheme 1.**
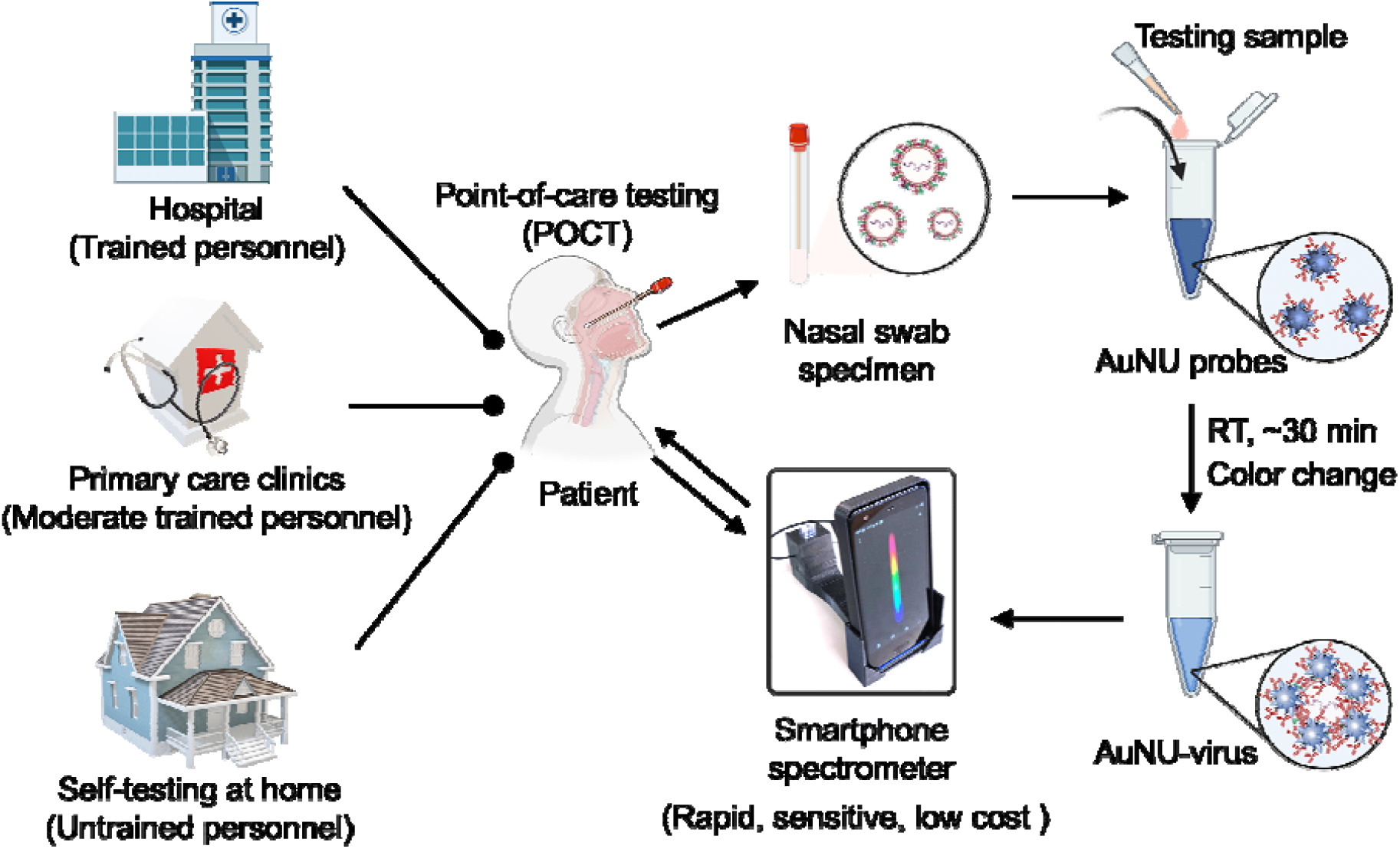
Schematic illustration of advances in Point-of-care testing (POCT) for a patient using the smartphone-based homogeneous assay.

## 2. Materials and methods

### 2.1. Materials

Tetrachloroauric (III) trihydrate (HAuCl_4_·3H_2_O, 16961-25-4, 99.9%), sodium citrate tribasic dihydrate (Na_3_CA·2H_2_O, 6132-04-3, ≥99%), and hydroquinone (123-31-9, ≥99%) were purchased from Sigma-aldrich. 3,3’-dithiobis (sulfosuccinimidyl propionate) (DTSSP, 21578, 50 mg) and borate buffer (1M, 28341) were purchased from Thermo Scientific. Gold nanorod (Citrate, GRCH800-1M) were purchased from nanoComposix, Inc. Amicon™ ultra centrifugal filter units (UFC510024) were purchased from Fisher Scientific. Palivizumab/Synagis (MedImmune, Gaithersburg, MD) was purchased from Children’s Health Retail Pharmacy. Bovine Serum Albumin (BSA) was purchased from BioPharm Laboratories, LLC. WarmStart® LAMP 2×Master Mix (M1700S) and LAMP Fluorescent dye (50×concentrate, B1700A) were purchased from New England Biolabs Inc. LAMP primer sequences were customized from Sigma-aldrich. RSV A2 quantitative genomic RNA (VR-1540DQ) was purchased from American Type Culture Collection (ATCC). Viral RNA Extraction Buffer (VRE100) was purchased from Sigma-aldrich. All aqueous solutions were prepared via ultra-pure (UP) water with a resistivity of 18.0 MΩ·cm.

### 2.2. Synthesis of spherical AuNP seeds

15 nm spherical AuNP seeds were synthesized using the Turkevich method with slight modifications.^15^ Briefly, 1 mL of HAuCl_4_·3H_2_O (25 mM) was added to 98mL of UP water and heated to boiling on the hot plate. While keeping vigorous stirring by magnetic bar, 1 mL of fresh prepared Na_3_CA·2H_2_O (112.2 mM) was injected into the boiled solution. The reaction was completed upon 10 minutes of heating and stirring. After cooling down to room temperature, the solution was stored in the dark for future use.

### 2.3. Size-controlled synthesis of Au nanourchins and Au nanospheres

Au nanourchins and Au nanospheres of different sizes were prepared based on a seed-mediated growth using 15 nm spherical AuNPs as seeds.^16-18^ Typically, the UP water, HAuCl_4_·3H_2_O, Na_3_CA·2H_2_O, and AuNPs seeds with a certain amount (**Table S2**) were added into a clean 250 mL Erlenmeyer flask orderly upon vigorous stirring at the room temperature. Then the hydroquinone was added subsequently. The reaction takes place immediately as indicated by the rapid color change. After stirring for over 2 hours, the final solution was kept in the dark for future use.

### 2.4. Antibody functionalization on AuNP probes for respiratory syncytial virus (RSV)

Synagis (Palivizumab) is a monoclonal antibody to RSV surface F protein^19^ and has been administrated for use in patients due to its significantly higher affinity and potency in neutralizing both A and B subtypes of RSV. We covalently conjugated Synagis onto plasmonic AuNPs with different shapes by DTSSP. The protocol was slightly revised based on previous work.^20^ Briefly, 5 mM DTSSP was mixed with Synagis and BSA in a molar ratio of 125:1, respectively. After 1 hour of incubation, the resulted DTSSP-Syn and DTSSP-BSA was dialyzed overnight in 2 mM borate buffer using 100 kDa molecular weight cut-off membrane at 4 °C. The resulted products were further purified using 10 kDa Amicon tubes. Then the DTSSP-Syn was incubated with AuNPs of various sizes and shapes for 2 hours on the ice bath to create plasmonic AuNP probes. Note that excess antibodies were used to cover and stabilize the AuNP probes entirely.^21^ We washed AuNP probes with 2 mM borate buffer by centrifugation at 4 °C, and then added 0.1% DTSSP-BSA into the solution with another 1-hour incubation. The backfilling process significantly alleviates the non-specific aggregation of AuNPs in a complex sample matrix.^22^ The final products were washed three times with 2mM borate buffer and stored in the 1.5 mL protein LoBind tube (Eppendorf 022431081).

### 2.5. Propagation and purification of RSV A2 strain

Human respiratory syncytial virus strain A2 (ATCC, Cat# VR-1540) was propagated by HEp-2 cells in 5% FBS/EMEM media (**Fig. S1**). Briefly, cells were inoculated with RSV at an MOI of 0.01 tissue culture infectious dose (TCID50)/cell. After 5-6 days of replication in cell culture, RSV A2 strain was purified from culture supernatant using 30%-60% (w/v) non-continuous sucrose density centrifugation (Beckman Coulter SW-28 rotor, 28000 rpm, 4°C for 90 minutes). Plaque titration by immunohistochemical staining technique (IHC) was used to determine the purified viral titers.^23^ Briefly, plaques formed on infected A549 cells were detected with a primary anti-RSV Fusion protein antibody (Palivizumab/Synagis, MedImmune, Gaithersburg, MD) and a secondary HRP conjugated anti-human antibody (Jackson Immuno Research Labs, Inc., West Grove, PA).

### 2.6. RSV detection by AuNP probes

The as-prepared AuNP probes were incubated with purified RSV and other close relative respiratory viruses (e.g., influenza type A virus (IAV), human metapneumovirus viruses (hMPV), and parainfluenza viruses (PIV)) as controls in a volume ratio of 2:1 (60 μL: 30 μL). The spectra were monitored after 30 minutes of incubation.

### 2.7. Digital Loop mediated-isothermal amplification (dLAMP) to quantify RSV A2 genomic RNA

LAMP primer sequences were designed using conserved regions of the matrix gene for RSV A.^24^ Detailed primer sequences were provided in **Table S3**. LAMP solution was prepared with a final volume of 20 µL containing 15 µL of isothermal master mix (including LAMP 2× Master Mix buffer, 10× primer mix, LAMP Fluorescent dye, and UP water with a volume ratio of 10:2:1:2) and 5 µL of target RNA. After gently mixing with a pipette, 15 µL of LAMP solution was then loaded onto QuantStudio™ digital chip and sealed by oil. The reaction was carried out at 65°C for 35 minutes, then followed by 2 minutes of heating at 95° to re-anneal the amplified DNA. Then the chips were observed under the fluorescent microscope.

### 2.8. RNA extraction from RSV A2

RSV A2 was lysed by viral RNA extraction buffer with a volume ratio of 3:1. After 5 minutes of incubation at room temperature, stabilized and exposed viral RNA was isolated using a Qiagen purification kit. Briefly, the binding buffer PB was added to the mixture of the extracts with a volume ratio of 5:1. Then the solution was transferred to RNA purification columns after mildly pipetting. Upon centrifugation at 17,000 ×g for 30 seconds, the flowthrough was disposed. Then 750 µL of washing buffer PE was added to each column. With 2^nd^ centrifugation at 17,000 ×g for 30 seconds, the RNA purification columns were transferred into clean 1.5 mL RNA Lobind tubes. Finally, 30-50 µL of elution buffer EB was added to each column and viral RNA was collected in the RNA Lobind tubes after 1 minute centrifugation at 17,000 ×g. We directly used dLAMP protocol described above for RNA detection from RSV A2.

### 2.9. Boundary element method to simulate plasmonic properties of AuNPs with different geometries

The plasmonic properties of AuNPs were calculated using the boundary element method (BEM) approach.^25^ Three models of AuNPs (i.e., spherical, rod, and urchin-like) were embedded in water as surrounding dielectric environment. The dielectric functions of gold used in the simulations were obtained from Johnson and Christy.^26^ All simulations were conducted via the MNPBEM toolbox in MATLAB (version 2021b).^27^

### 2.10. Smartphone spectrometer device

The smartphone spectrometer was assembled using a low-cost HTC U11 smartphone (1/2.55’’ sensor size). The fabrication process was optimized and referred to a previous work.^28^ Briefly, the rear-face camera was used as an array detector. A custom-designed cradle with a diffraction grating holder, cuvette holder, and two plastic capillaries have been fabricated by a 3D printer using Acrylonitrile Butadiene Styrene (ABS) polymer. One of the plastic fibers (1.5 mm) carries the phone back flashlight to the cuvette holder, and another fiber collects transmitted light (0.25 mm) to the linear diffraction grating slide (Amazon, 300-700 nm, 1000 lines/mm). Incoming light hits the diffraction grating with ∼42° to the normal, and first-order diffracted light falls on top of the phone camera. Three lasers with different wavelengths (405, 532, and 641 nm) were used to calibrate the pixel index and the corresponding wavelength. The conversion factor adapted from the linear relationship between wavelength and pixel index was 0.293 nm/pixel. The spectrophotograms are collected as RAW image format and processed by customized MATLAB script. The RGB information from RAW images were converted to the Hue–Saturation–Value (HSV), where the value (V) represents the light transmission through the solution. The absorbance intensity (*A*) was calculated based on Beer–Lambert law:

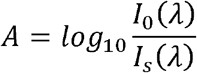

where *I*_*0*_ is the transmitted light intensity of the blank control solution (i.e., 2 mM borate buffer in this study), and *I*_*s*_ is the transmitted light intensity of the testing solution.

### 2.11. Measurements and characterization

The size distribution of nanoparticles was measured by dynamic light scattering (Malvern ZetaSizer Nano ZS). The absorbance of sample solutions was read by microplate reader (Synergy 2, BioTek). Extinction spectra was obtained using a spectrophotometer (DU800, Beckman Coulter). The transmission electron microscopy (TEM) images were taken using a JEOL JEM-2010 microscope.

### 2.12. Biosafety statements

The research project was approved and performed strictly in adherence to CDC/NIH guidelines. The experimental protocols and the use of human-related clinical specimens were approved by Institutional Biosafety & Chemical Safety Committee (IBCC, #190517) and the Institutional Review Board (IRB, #MR15-189) from the University of Texas at Dallas.

## 3. Results and discussions

### 3.1. Au nanourchin with long protrusions provides the most sensitive detection among three different nanoparticle shapes

First, we compared the analytical performance for three nanoparticle shapes for colorimetric detection of intact viruses. Nanoparticles clustering around the virus leads to a color change due to plasmonic interactions between the nanoparticles (**Fig. 1A**). We prepared Au nanourchins (AuNUs), Au nanospheres (AuNSs), and Au nanorods (AuNRs, nanoComposix) and characterized the nanoparticles with transmission electron microscopy (TEM, **Fig. 1B-D**). **Fig. S2A-C** show their corresponding sizes to be 75 nm ± 6 nm (defined as tip-to-tip length of an urchin), 50 nm ±5 nm (diameter of spheres), and 50 nm±7 nm and 15 nm ± 2 nm (length and width of rods). All the AuNPs have citrate ions as capping ligands on the surface that facilitates the subsequent antibody (i.e., Synagis/Palivizumab) conjugation via a covalent crosslinker DTSSP (**Fig. S2D**). The Synagis as a passive immunoprophylactic agent and specifically targets the fusion (F) proteins on the RSV surface.^29^ Synagis coating on AuNPs was verified by the increase of hydrodynamic sizes (**Fig. S2E-G**). Then the Synagis-modified AuNPs were ready for the RSV detection. Here, all AuNP probes with the same optical density (O.D. = 5) were incubated with serial dilutions of RSV stocks at room temperature for 30 minutes and measured by UV-Vis spectrometer (**Fig. 1E-G**). The spectra show an obvious drop in the peak intensity for all cases with higher RSV concentration. We then applied a ratiometric method to quantify the results. Taking AuNUs as an example, we generated the calibration curve by plotting the absorbance intensity (*I*_*Abs*_) ratio of 800 nm to 660 nm against the viral titers (**Fig. 1H**). A linear relationship (R^2^ = 0.99) was observed in the range of 2,500-50,000 PFU/mL (**Fig. 1H inset**). The limit of detection (LOD) was determined to be 1,402 PFU/mL by calculating the concentration corresponding to a signal that is 3 times the standard deviation above the blank control calibrator.^30^ The same strategy was applied to other AuNP probes for the LOD calculation (**Fig. 1F** for AuNSs and **Fig. 1G** for AuNRs). Their corresponding linear detection ranges and LOD were determined to be 10,000-200,000 PFU/mL and 8,000 PFU/mL, and 10,000-100,000 PFU/mL and 4,360 PFU/mL, respectively. AuNUs provided the most sensitive detection for RSV, which was about 5.7- and 3.1-times lower than that of AuNSs and AuNRs.

**Fig. 1.**
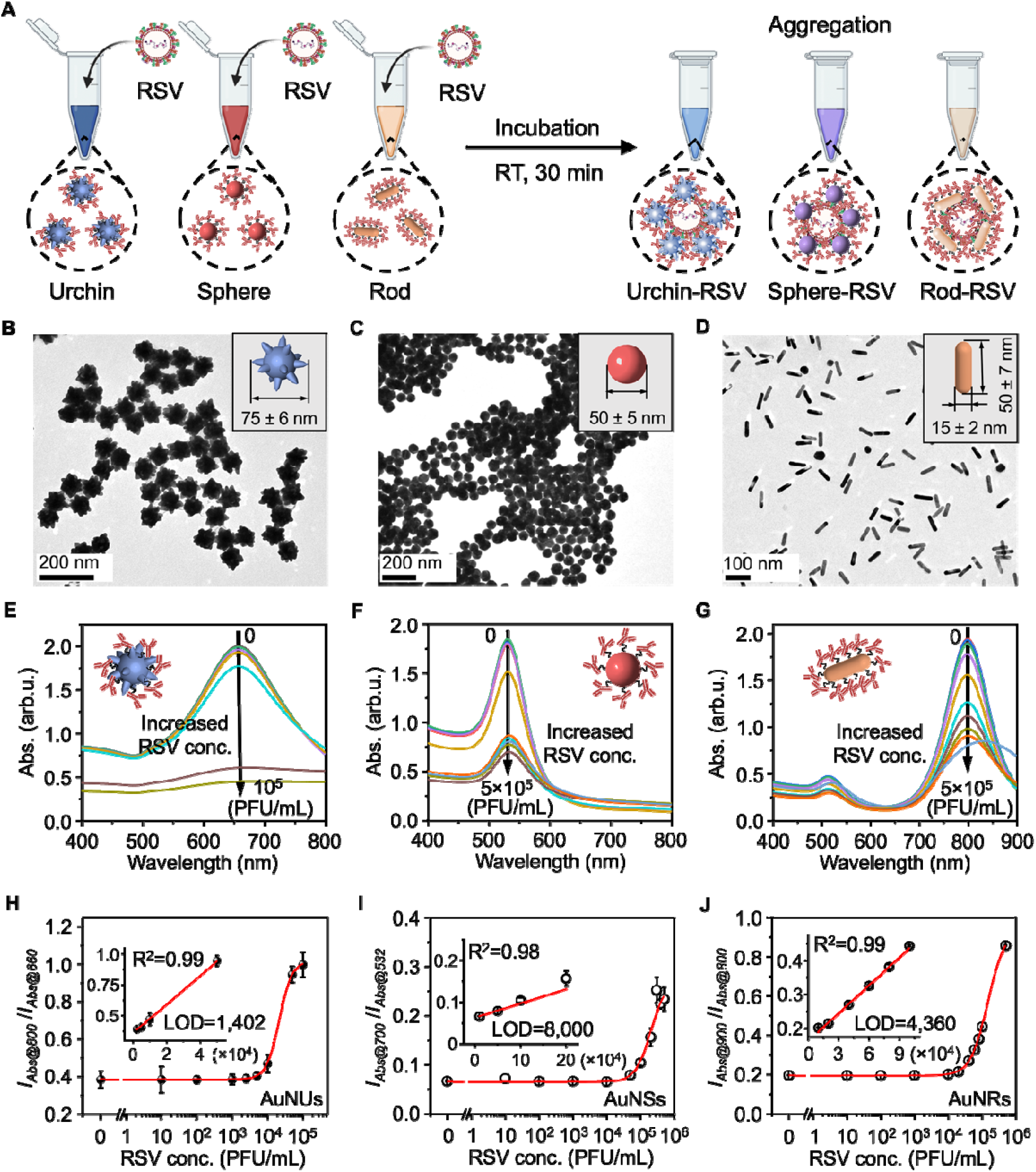
Respiratory syncytial virus (RSV) detection using plasmonic AuNPs with different shapes. (A) Schematic illustration of AuNPs aggregation induced colorimetric detection of viruses. (B-D) Transmission electron microscopy (TEM) image of (B) Au nanourchins (AuNUs), (C) Au nanospheres (AuNSs) and (D) Au nanorods (AuNRs). Insets show the measured size for each NP. (E-G) Colorimetric detection using (E) AuNUs, (F) AuNSs, and (G) AuNRs as probes. Each inset shows the model of the probe for detection of RSV with serial dilutions (0-100,000 PFU/mL for AuNUs, and 0-500,000 PFU/mL for AuNSs and AuNRs). (H-J) Corresponding calibration curves by plotting the absorbance intensity (*I*_*Abs*_) ratio at two wavelengths against RSV titers. The error bars indicate the standard deviations (*n* = 3). Insets show the linear detection range and calculated limit of detection (LOD).

To verify that the assay is specific for RSV detection, we incubated the AuNP probes with other closely related respiratory viruses. These include influenza type A virus (IAV), human metapneumovirus (hMPV), and parainfluenza viruses (PIV) (**Fig. 2A-C**). Spectral analysis suggests that only the RSV leads to the peak intensity decrease (**Fig. 2D**) and confirms the as-prepared AuNP probes have good detection specificity among those respiratory viruses.

**Fig. 2.**
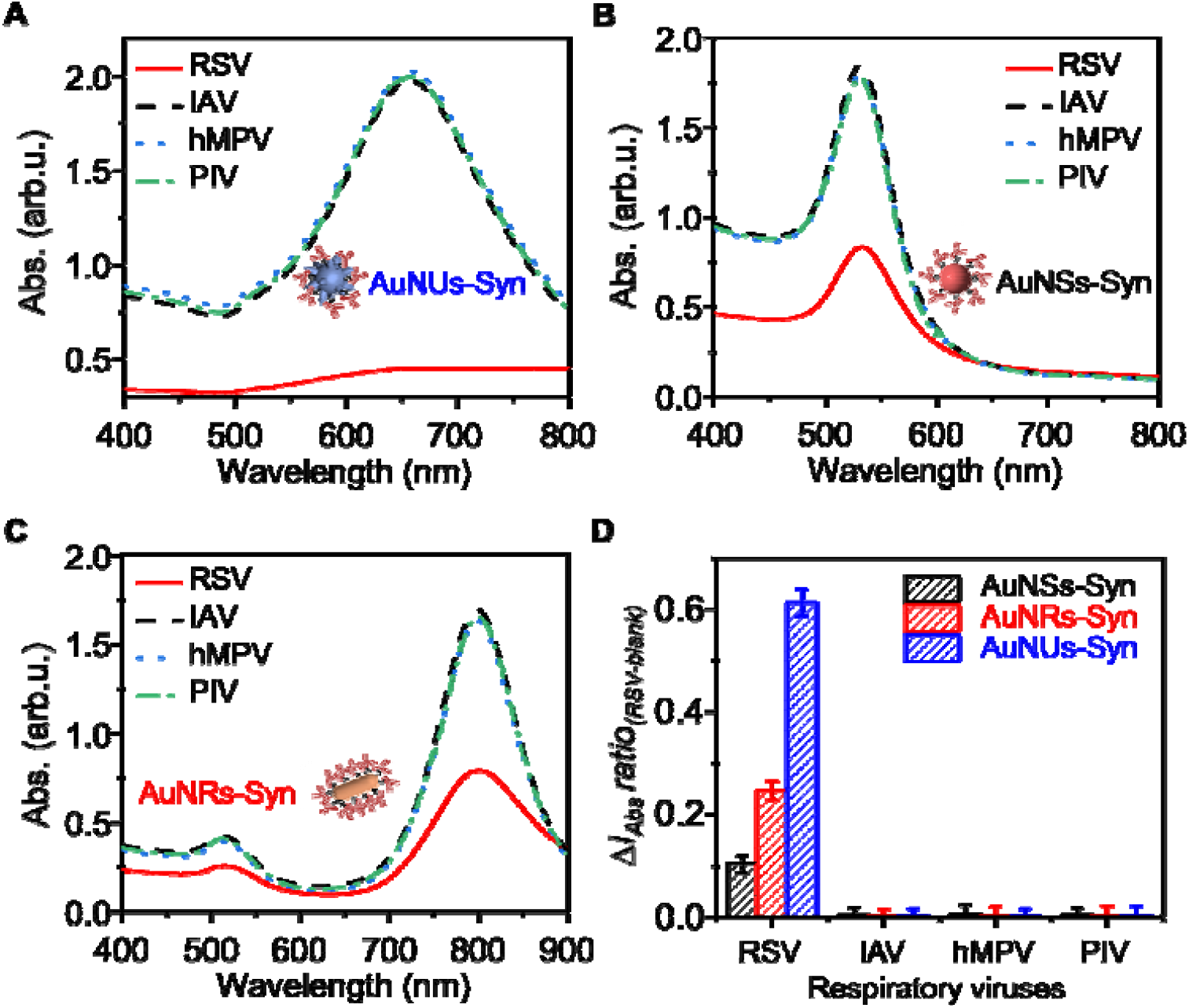
Selective detection of RSV among closely relative respiratory viruses. (A-C) Spectral measurements of RSV detection using AuNUs, AuNSs, and AuNRs, respectively. (D) The change of extinction ratio obtained from corresponding calibration curves demonstrate that all the AuNP probes allows selective detection of RSV at 10^5^ PFU/mL from other respiratory viruses including influenza type A virus (IAV), human metapneumovirus (hMPV), parainfluenza viruses (PIV). The error bars indicate the standard deviations (*n* = 3).

To fully understand the impacts of AuNPs’ physical properties on the assay performance, we further evaluated the effect of AuNPs size and concentration on RSV detection. Since the size of the urchin-like and spherical NPs can be readily tuned, we chose them as a model to investigate the effect of particle size. By adjusting the amount of the reagents in the reaction, we obtained AuNUs with different tip lengths (65 nm and 75 nm, **Fig. S3**). The same method was adapted to the size-controllable synthesis of AuNSs. Then we incubated serial dilutions of RSV with AuNU (**Fig. S4A-B**) and AuNS probes (**Fig. S4C-F**) of different sizes and kept them at the same concentration (5 O.D.). In addition to 75 nm AuNU and 50 nm AuNSs in previous section (**Fig. 1E-F** and **H-I**, LOD = 1,402 PFU/mL and 8,000 PFU/mL), **Fig. S5** shows that the LODs for 65 nm AuNUs, 15 nm AuNSs, and 100 nm AuNSs were 7,999, 34,000, and 8,700 PFU/mL, respectively. Smaller size AuNUs result in less sensitive detection (7,999 PFU/mL versus 1,402 PFU/mL). For spherical NPs, increasing the AuNSs size from 15 nm to 50 nm improves the detection sensitivity (4.25-fold enhancement), while further increasing the size to 100 nm leads to less sensitive detection (8,000 PFU/mL versus 8,700 PFU/mL).

We then performed parallel experiments using AuNPs (i.e., AuNUs, AuNSs, and AuNRs) of different optical densities (1, 5, and 10 O.D.) to assess the effect of NP’s concentration on assay sensitivity (**Fig. S6**). **Fig. S7** shows that increasing the particle concentration from 1 O.D. to 5 O.D. leads to lower LOD values (better detection sensitivity) for all cases. When further increasing the NP’s concentration to 10 O.D., a U-shaped dependence of the LOD on the particle concentration was observed, indicating that there is a critical particle concentration in favor of the most sensitive detection. Notably, AuNP probes with 5 O.D. are sufficient to provide sensitive detection for RSV. This phenomenon has also been reported for the nucleic acid detection.^31^ Among all the cases tested, AuNUs with longer protrusions show the lowest LOD at 1,402 PFU/mL.

To convert the plaque-forming unit (PFU) into RNA copies, we performed the digital loop-mediated amplification (dLAMP) and quantified the RSV titers. **Fig. S8A-H** show the dLAMP images using quantitative genomic RSV A2 RNA as standards. A calibration curve was plotted by counting the fraction of positively fluorescent wells (*f*_on_) over the total wells against the RNA concentrations (**Fig. S8I**). The LOD is calculated at 2 copies/µL. We then lysed serial dilutions of RSV samples and purified the resulted viral RNA before taking them into the dLAMP reactions. **Fig. S9** show the corresponding detection results. The LOD is calculated to be 141 PFU/mL. Based on the *f*_on_ values, we can correlate RSV titers (PFU/mL) and RNA concentrations (copies/µL, **Fig. S10**), suggesting that 100 PFU/mL is approximately 1.2 copies/µL. Then we calculated the detection limit of RSV by 75 nm AuNUs as 17 copies/µL. We summarized our findings in **Table 1** for the three nanoparticle shapes with various sizes and concentrations. The detection sensitivity is competitive with some molecular tests (LOD = 10-100 copies/µL).^32^ Therefore, the AuNU provides a highly sensitive and specific detection for intact virus with a one-step homogeneous assay.

**Table 1.** Comparison of RSV detection performance for three different nanoparticle geometries (sphere, rod, urchin) and concentrations (1, 5, 10 OD). Testing volume = 30 µL.

| **AuNP shape** | **Dimension (nm)** | **Concentration (O.D.)** | **LOD (PFU/mL)** | **LOD (copies/μL)** | **LOD (PFU)** |
| --- | --- | --- | --- | --- | --- |
| Sphere | 15 | 1 | 280,000 | 3,360 | 8400 |
|  |  | 5 | 34,000 | 408 | 1020 |
|  |  | 10 | 200,000 | 2,400 | 6000 |
|  | 50 | 1 | 29,000 | 348 | 870 |
|  |  | 5 | 8,000 | 96 | 240 |
|  |  | 10 | 7,900 | 95 | 237 |
|  | 100 | 1 | 14,000 | 168 | 420 |
|  |  | 5 | 8,700 | 104 | 261 |
|  |  | 10 | 9,600 | 115 | 288 |
| Rod | 50 (length)<br>×<br>15 (width) | 1 | 8,330 | 100 | 250 |
|  |  | 5 | 4,360 | 52 | 131 |
|  |  | 10 | 5,129 | 62 | 154 |
| Urchin | 65 nm | 1 | 16,000 | 192 | 480 |
|  |  | 5 | 7,999 | 96 | 240 |
|  |  | 10 | 7,300 | 88 | 219 |
|  | 75 nm | 1 | 2,058 | 25 | 62 |
|  |  | 5 | **1,402** | **17** | **42** |
|  |  | 10 | 1,543 | 19 | 46 |

### 3.2. Electron microscopy imaging reveals high virus binding capability for AuNU with longer protrusions and AuNR

Second, we examined the extent of NPs accumulation on the virus surface due to its critical role in the colorimetric assay. We took TEM images from completed assay solutions using AuNP probes with different shapes, sizes, and concentrations (**Fig. 3A**) under the same virus titer (10^6^ PFU/mL). The virions show different morphologies^33^ in the negative stained images (highlighted by red arrows). An important observation is that the AuNUs with longer protrusions and AuNRs show much more dense particle coverage on virions than AuNSs. For a quantitative comparison, we measured the particle coverage on the virion-probes complexes and determine the binding effectiveness (BE, defined as the area of the viruses covered by the NPs in the 2D image). **Fig. 3B** shows the BE results for all AuNP probes and confirms that 75 nm AuNU and AuNR overall have higher binding to the viruses at different nanoparticle concentrations (1, 5, and 10 O.D.). The particle coverage on virions increases with higher nanoparticle concentrations (1 vs 5 O.D), while the coverage saturates for AuNRs and AuNUs when further increasing the concentration from 5 to 10 O.D. Furthermore, we also observed more free NPs in the cases of high AuNP concentrations (e.g., 10 O.D. AuNSs and AuNRs), consistent with the increased background color in the assay. The TEM analysis suggests that non-spherical nanoparticles with higher surface area, either by rod shape or long protrusions on the nanourchin surface, increase the virus binding capability. The high virus-binding capability for AuNRs and AuNUs reveals a key mechanism for the high detection sensitivity with these nanoparticles.

**Fig. 3.**
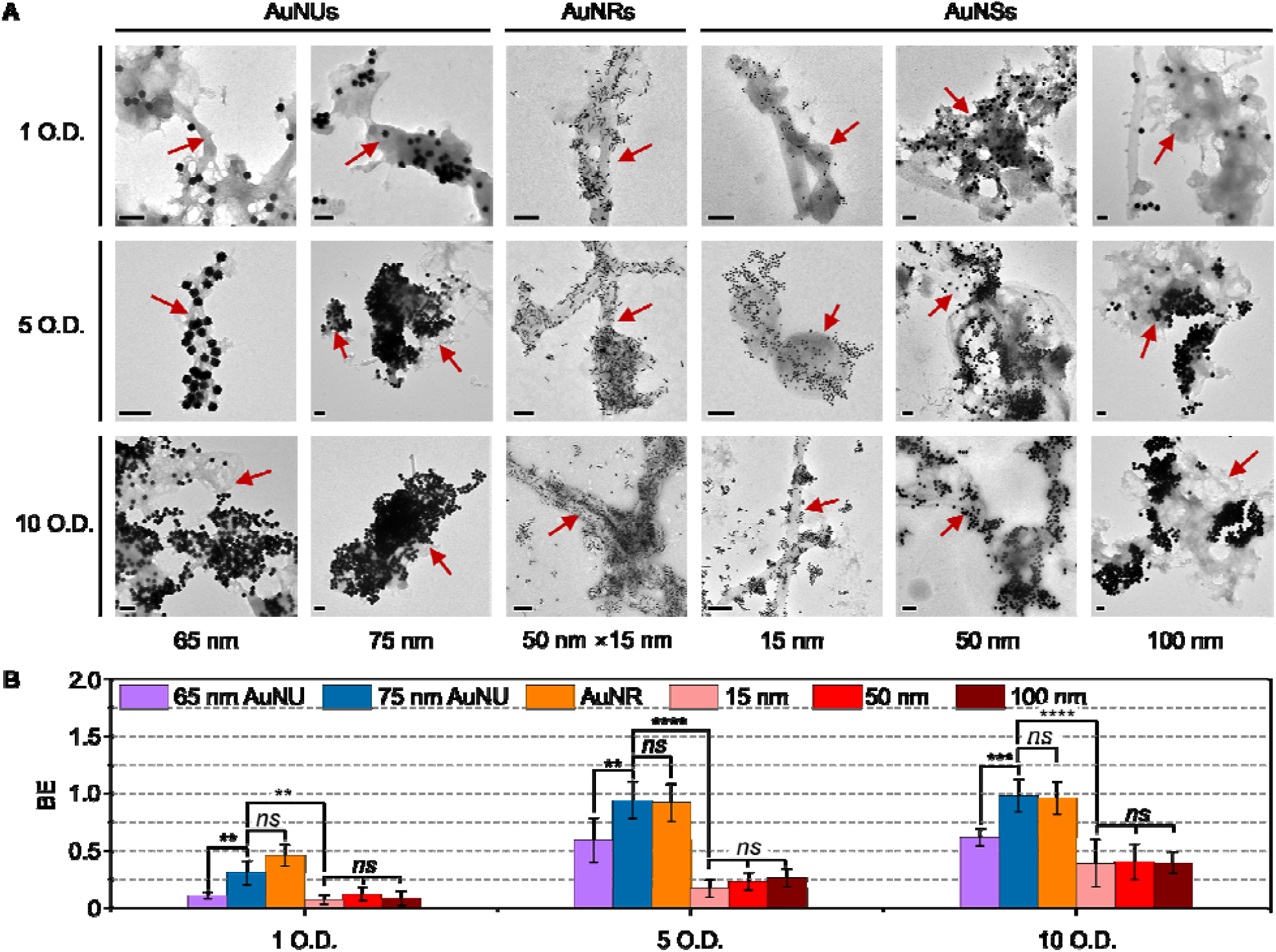
AuNUs with longer protrusions and AuNRs show higher virus binding effectiveness (BE) than AuNSs. (A) TEM images of AuNUs, AuNRs, and AuNSs probes with different optical density (O.D.) after incubating with RSV at 10^6^ PFU/mL. Scale bar = 200 nm. Red arrows indicate negative stained RSV A2 strains via 2% uranyl acetate. Note that RSV is variable in shape and size (e.g., spherical, or filamentous). (B) Binding effectiveness (BE) analysis for different AuNP probes. BE is defined as the area of the viruses covered by the NPs in the 2D image. NPs counting and area measurements were performed by Image J. The total number of TEM images used to analyze the AuNP probes at each concentration is *n*=6 from 3 samples. ns, P > 0.05; **, P < 0.01; ****, P < 0.0001.

### 3.3. Modeling the plasmonic coupling shows that AuNU with longer protrusions has strongest interaction at long distances (10 nm)

Third, we investigated the plasmonic coupling strength for the nanoparticles, another key factor in determining the colorimetric assay performance, with computational simulations. Among different computational techniques (e.g., Mie theory,^34^ discrete dipole approximation,^35, 36^ finite element method,^37^ finite difference time domain^38^), boundary element method (BEM) provides simple and fast simulation for AuNPs with complex geometries. Particularly, we calculated the extinction cross-section area as a metric for the AuNP plasmonic coupling, as colorimetric assays essentially measure the extinction or absorbance change of the AuNPs probes. For simplicity, we calculated the plasmonic coupling strength between two coupled AuNPs. We first built a model of two identical AuNSs with interparticle distance *d* of 0.1-10 nm (**Fig. 4A**). **Fig. S11** show the simulated extinction cross-section for coupled AuNSs with different sizes. Decreasing the interparticle distance leads to a red shift in the plasmonic peak (to longer wavelength) and an increase in the extinction cross-section. Different particle sizes show the same trend. Furthermore, the larger AuNSs have a greater peak shift than smaller ones (e.g., 15 nm). **Fig. 4B-C** show the change of extinction cross-section (ΔExt_@SPR_) and the peak shift of coupled AuNSs against that of a single particle. Clearly, both values of ΔExt_@SPR_ and peak shift decrease as the interparticle distance (*d*) increases for a given particle size. While fixing *d* (< 5 nm), larger particle size has higher ΔExt_@SPR_ and peak shift. Notably, for *d* > 5 nm, the ΔExt_@SPR_ becomes close for 50 nm and 100 nm AuNSs. This suggests that further increasing AuNP size does not necessarily improve the plasmonic coupling.

**Fig. 4.**
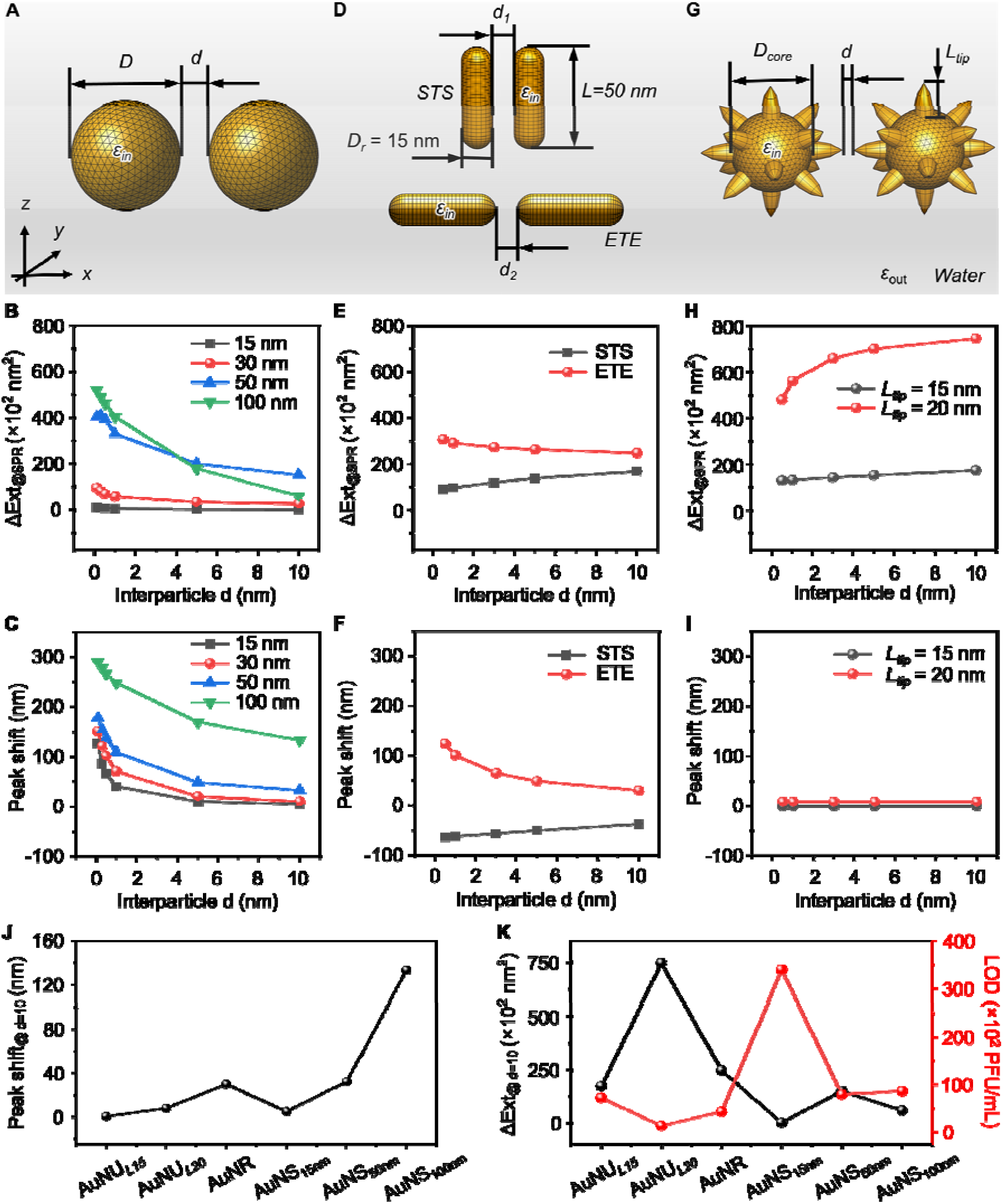
BEM simulation of plasmonic coupling properties for AuNP probes with different shapes. Models for two coupled (A) AuNSs, (D) AuNRs, and (G) AuNUs. The calculated change of extinction cross-section (ΔExt_@SPR_) and peak shift for coupled AuNSs (B, C), AuNRs with side-to-side (STS) and end-to-end (ETE) orientations (E, F), and AuNUs (H, I) as a function of interparticle distance *d*. The extinction cross-section and the peak position of a single AuNP were used as a reference. (J-K) Comparison of peak shift and extinction cross-section change (ΔExt_@SPR_) for AuNP probes at *d* = 10 nm.

We then examined a model for coupled AuNRs with dimension of 50 nm × 15 nm. Because of their anisotropic shape, different orientations of the AuNRs lead to various modes of coupling. For simplicity, we built two representative models (**Fig. 4D**) to calculate their plasmonic coupling strength, i.e., side-to-side (STS) and edge-to-edge (ETE). **Fig. S12** shows the simulation results of extinction cross-section with STS and ETE coupling. Three linear polarization directions (i.e., x-, y-, and z-directions) were considered in the simulations since the optical properties of AuNRs are polarization-dependent (**Fig. S13 and S14**). The two coupling modes lead to opposite peak shift, where a blue shift of longitudinal peak occurs for the STS and a red shift for ETE, consistent with previous work.^39^ Increasing the inter-particle distance (*d*) leads to an increase of ΔExt_@SPR_ and peak shift for STS, and a decrease of ΔExt_@SPR_ and peak shift for ETE (**Fig. 4E-F**). Interestingly, ETE mode has higher ΔExt_@SPR_ values than the STS mode, regardless of the coupling gap.

Next, we examined for AuNUs which have irregular tips on the surface. For simplicity, we constructed the model with a core-tips structure and evaluated the influence of a set of parameters on the optical response, including tip number (*n*), tip length (*L*), and core size (*D*) based on the TEM (**Fig. S15A**). We first analyzed the model with *D* = 50 nm and *L* = 20 nm for different tip numbers (*n* = 1, 6, and 14). **Fig. S15B** shows the simulation results of the extinction cross-section for individual AuNUs. It is worth mentioning that the simulated spectra of AuNUs can be assumed as the main plasmon mode confined on the tips (LSPR at around 700 nm) and a secondary mode confined within the core (LSPR at around 520 nm).^40, 41^ We found that additional tips on the surface increase the extinction cross-section at the longitudinal resonance (∼700 nm) and minor blue shift of the peak. Moreover, the appearance of multiple peaks can be observed as the shape of the AuNUs becomes more complicated. On the other hand, increasing the tip length *L* (**Fig. S15C**) or core size *D* (**Fig. S15D**) of an AuNU with a fixed tip number (*n* = 14) leads to the increase of extinction cross-section as well. Detailed polarization-dependent plots of each case are shown in **Fig. S16-S18. Fig. S19** shows the simulated extinction cross-section at different interparticle distances for coupled AuNUs (tip-to-tip, *D* = 50 nm, *L* = 15 nm or 20 nm, and *n* = 14). Interestingly, we found that the magnitude of the extinction cross-section (ΔExt_@SPR_) changes with the interparticle distance while there is minimal peak shift regardless of the interparticle distance between two coupled AuNUs (**Fig. 4G-I**). Longer tips lead to a larger change in the extinction cross-section (ΔExt_@SPR_), and therefore plasmonic coupling strength.

Other factors such as the binding orientation among anisotropic nanoprobes can influence plasmonic coupling. For simplicity, we again calculated the plasmonic coupling strength for two anisotropic NPs (i.e., AuNRs and AuNUs) with different coupling angles (θ). The interparticle distance (*d*) was set at 10 nm since it represents the closest interparticle distance in the case of antibodies conjugation on the nanoparticle surface (e.g., the size of antibodies is 5∼10 nm).^42^ **Fig. S20A, B** show the models and corresponding simulation results for AuNRs with various θ. When altering the θ from 0° to 180°, the rod dimer shows anti-bonding and bonding resonance at about 700 nm and 750 nm, which agrees with previous studies.^43-46^ Results suggest that θ = 180° as the ETE coupling case provides the highest plasmonic coupling strength. In the case of AuNUs (**Fig. S20C, D)**, it can be seen that AuNUs show magnitude changes in extinction cross-section without obvious peak shifts as θ changes. While there is some orientation dependence, the overall trends of the extinction cross-section for both AuNRs and AuNUs are in the same range as the previous simple model (**Fig. 4**).

To better understand the effect of the plasmonic coupling mechanism on colorimetric sensing performance, we compared the simulation results at *d* = 10 nm for all AuNPs employed for RSV detection. **Fig. 4J-K** suggest that ΔExt_@SPR_ is a more important factor to evaluate the detection performance than that of the peak shift. Non-spherical AuNP probes with higher values of ΔExt_@SPR_ tend to provide better detection sensitivity, where AuNUs_*L*20_ (longer tip length, *L* = 20 nm) show 4.8- and 3-times higher ΔExt_@SPR_ than that of AuNSs (50 nm) and AuNRs, respectively. Our results suggest that the extinction cross-section change (ΔExt_@SPR_) is a good indicator for the plasmonic coupling strength and assay performance.

### 3.4. Smartphone spectrometer enables point-of-care diagnostics of clinical samples with similar sensitivity to laboratory spectrometer

We further fabricated a 3D-printed device that can be integrated with a smartphone for virus detection at the point of care (**Fig. 5A**). Specifically, the device prototype includes a phone case to slide the low-cost smartphone (Android, HTC U11, sensor size 1/2.55’’), a compartment that holds a cuvette (**Fig. S21**) for testing and blocks out environmental light, and a bundle of plastic fibers for light propagation. The integrated device has a single diffraction grating to divide the light and contains no electronic components in a low-cost and simple design. For colorimetric measurements, we simply apply the flashlight from the smartphone and take the RGB images. This facilitates sample testing in a “plug and play” fashion. The pixel index from RAW images was converted to wavelength by three lasers of different wavelengths (405, 532, and 641 nm, **Fig. 5B**).

**Fig. 5.**
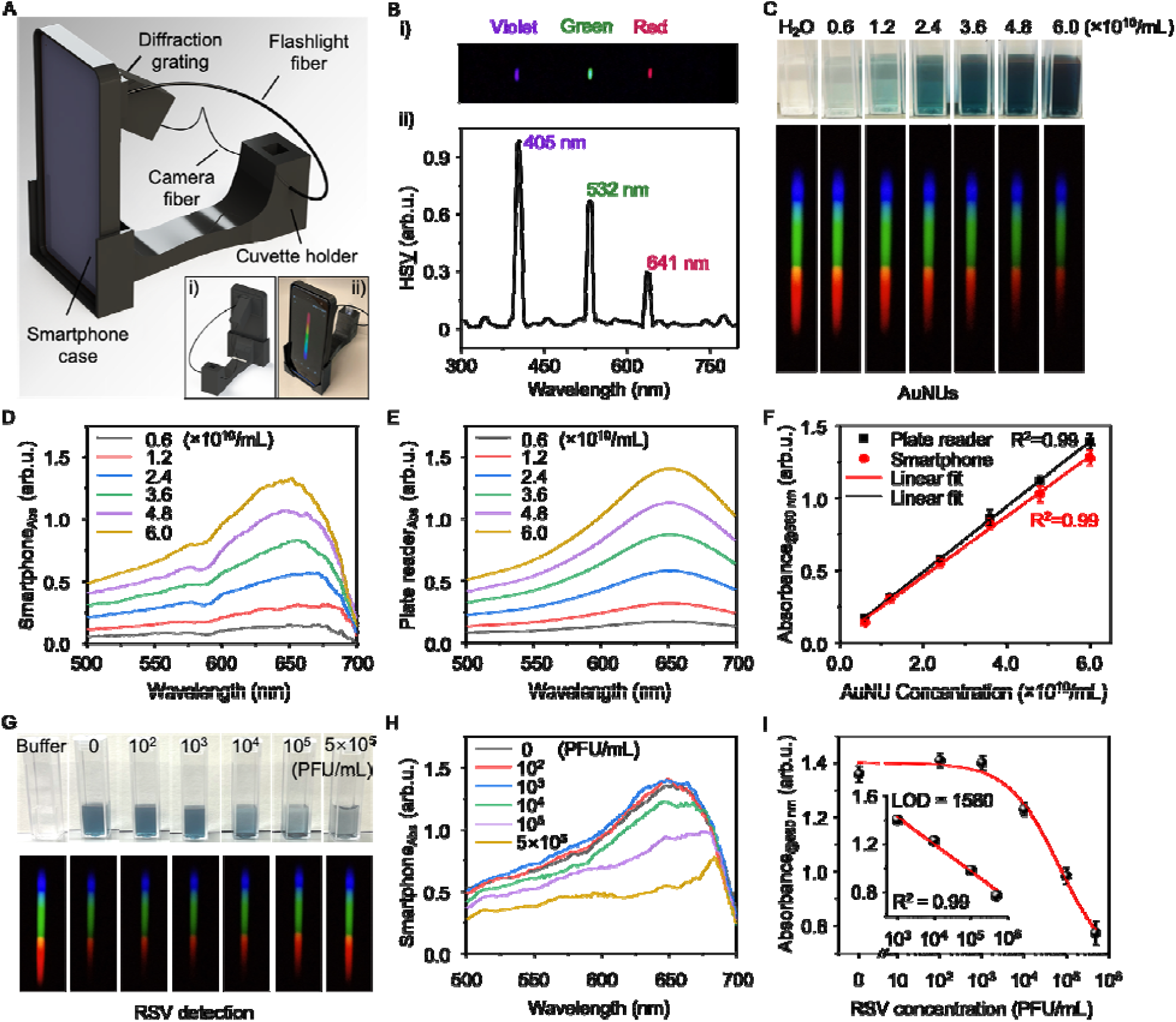
Smartphone-based spectrometer enables point-of-care colorimetric detection. (A) CAD drawing of the smartphone spectrometer. Insets show the backside of the design (i) and a photograph of the assembled smartphone spectrometer (ii). (B) Pixel index and wavelength calibration by three lasers. (i) The spectral images obtained by the plastic fibers from violet (405nm), green (532 nm), red (641 nm) lasers. (ii) The output spectrum was extracted from th image after calibration. HS<u>V</u> means the value in Hue-saturation-value (HSV) was plotted. (C) Serial dilutions of AuNU suspensions were imaged by smartphone spectrometer. (D-E) Absorbance spectrum obtained from smartphone (D) and commercial microplate reader (E) for serial dilutions of AuNU suspensions. (F) Comparison of the measurement performance of the smartphone spectrometer and the commercial microplate reader. The error bars indicate the standard deviations (*n* = 3). (G-I) RSV detection by the smartphone device. (G) The photographs and spectral images for RSV detection. (H) Corresponding absorbance spectrum were extracted from (G). (I) Calibration curves by plotting the peak absorbance at 660 nm against RSV titers. The error bars indicate the standard deviations (*n* = 3). Inset shows the linear detection range and calculated limit of detection (LOD) for AuNUs using smartphone spectrometer.

To validate the smartphone-based colorimetric reader, we measured serial dilutions of 75 nm AuNU suspensions (**Fig. 5C**) and compared the results to a commercial microplate reader. **Fig. 5D** and **Fig. 5E** show the absorbance spectra obtained from a smartphone and a commercial microplate reader, respectively. The smartphone measurements show diminishing values at the upper limit of the diffraction grating (∼700 nm) but give similar peak values with the microplate reader. Herein we plotted the peak absorbance at 660 nm against the particle concentrations (**Fig. 5F**). Both devices showed similar detection results and confirmed the utility of smartphone-based reader for colorimetric measurements. Moving forward, we used this device to test completed assay solutions containing 75 nm AuNU probes and serial dilutions of RSV. We analyzed the smartphone data using the peak absorbance instead of the previous ratiometric method since the smartphone measurements are not accurate when approaching 700 nm. Also, AuNU probes mainly show intensity change and minimal peak shift in virus samples, as shown in previous experiments (**Figure 1E**). Therefore, the peak absorbance values are preferred to quantify the limit of detection. **Fig. 5G-I** show that the detection limit was calculated to be ∼ 1580 PFU/mL, similar to the microplate reader result (∼1,402 PFU/mL).

Lastly, we explored the potential clinical application of infectious disease diagnostics using the smartphone-based spectrometer. We collected nasal swab samples from healthy adults and spiked RSV of different concentrations into the nasal swab sample solutions. We directly tested those samples with 75 nm AuNU probes and measured them by smartphone spectrometer. Our result suggests sensitive detection in the complex sample matrix with high accuracy (**Table 2**), demonstrating the feasibility of mobile detection at the point of care.

**Table 2.** Analytical performance of the smartphone-based diagnosis of RSV-spiked nasal swab samples.

| **Spiked RSV**<br>**(PFU/mL)** | **Estimated viral RNA**<br>**(copies/μL)** | **Positive test #** | **Total test #** | **Accuracy** |
| --- | --- | --- | --- | --- |
| 0 | 0 | 0 | 5 | 100% |
| 2,000 | 24 | 4 | 5 | 80% |
| 10,000 | 120 | 5 | 5 | 100% |
| 100,000 | 1200 | 5 | 5 | 100% |

## 4. Conclusions

In summary, we have developed a rapid and sensitive PCA for RSV detection. We demonstrated that AuNUs with long protrusions achieved the lowest detection limit among AuNRs and AuNSs. The improved analytical performance of AuNUs can be attributed to higher virus binding and stronger plasmonic coupling at long distances. We further assembled a 3D-printed smartphone spectrometer for portable and colorimetric measurements, allowing for sample-to-answer and quantitative tests. Therefore, the AuNU-based PCA has the great potential to be employed for large-scale tests and to combat infectious disease spread.

## Data Availability

All data produced in the present work are contained in the manuscript

## Associated content

### Supporting information

The supporting information was submitted along with the main text.

## Author information

### Corresponding Author

*

### Author contributions

Y.L. and H.Y. carried out the AuNSs and AuNUs experiments and performed the data analysis. Y.L. and A.B. fabricated the smartphone spectrometer and developed codes for image analysis. Y.L., T.Z., and S.A.M.A.P. conducted the AuNRs experiments and implemented the data analysis. Y.L. and Q.C. synthesized and characterized the AuNUs. Y.L. and H.H. prepared the virus samples. Y.L. collected and analyzed TEM images. Y.L. and H.Y. performed the dLAMP assay. Y.L. and X.C. analyzed dLAMP images. Y.L. performed the BEM simulation. Y.L., H.Y., and Z.Q. wrote the manuscript. Y.L. and Z.Q. conceived the original idea and supervised the project. All authors revised the manuscript and have given approval to the final version of the manuscript.

## Funding sources

Research reported in this manuscript was partially supported by National Institutes of Health (NIH) grants R21AI140462 and R01AI151374, and U.S. Department of Defense (DOD) grant W81XWH-20-1-0106 to Z.Q.

## Notes

Y.L. Z.Q. H.Y. and J.S.K. are the inventors of a provisional patent related to this work filed by the University of Texas at Dallas. Z.Q. and J.S.K. hold equity interest in Avsana Labs, Incorporated, which aims to commercialize the technology. The remaining authors declare no competing financial interest.

## Notes

### Competing Interest Statement

Y.L. Z.Q. H.Y. and J.S.K. are the inventors on a provisional patent related to this work filed by the University of Texas at Dallas. Z.Q. and J.S.K. hold equity interest in Avsana Labs, Incorporated, which aims to commercialize the technology. The remaining authors declare no competing financial interest.

### Author Declarations

The University of Texas at Dallas Institutional Biosafety & Chemical Safety Committee and the Institutional Review Board gave ethical approval for this work.

