## Supplemental Information for "Gold Nanourchins Improve Virus Targeting and Plasmonic Coupling for Virus Diagnosis on a Smartphone Platform"

**Table of Contents:**

**Table S1** Antigen diagnostics for pathogens using plasmonic gold nanoparticles (NPs).

**Table S2** Reagents for size-controlled synthesis of Au nanourchins and Au nanospheres.

**Table S3** RSV A2 primer sequences for LAMP Assay.

**Fig. S1** Large scale propagation of Respiratory syncytial virus (RSV) A2 stain.

**Fig. S2** Characterization of AuNP probes.

**Fig. S3** Synthesis and characterization of AuNUs of different sizes.

**Fig. S4** Characterization of 65 nm AuNUs with short tips and different sizes of AuNSs.

**Fig. S5** Respiratory syncytial virus (RSV) detection using plasmonic AuNUs and AuNSs with different sizes.

**Fig. S6** Respiratory syncytial virus (RSV) detection using AuNP probes of different concentration.

**Fig. S7** Effect of AuNP probes concentration on RSV detection. O.D.= Optical density.

**Fig. S8** dLAMP of quantitative genomic RNA for RSV A2 strain.

**Fig. S9** dLAMP assay for detection of RNA extracts from RSV A2 strain.

**Fig. S10** Calibration of RSV A2 and corresponding RNA concentration by dLAMP assay.

**Fig. S11** BEM simulation of the plasmonic coupling properties for AuNSs.

**Fig. S12** BEM simulation of the plasmonic coupling properties for AuNRs.

**Fig. S13** BEM simulation of the extinction cross-section for STS-coupled AuNRs with x-, y-, and z- polarization directions.

**Fig. S14** BEM simulation of the extinction cross-section for ETE-coupled AuNRs with x-, y-, and z- polarization directions.

**Fig. S15** BEM simulation of plasmonic properties of AuNUs.

**Fig. S16** BEM simulation of the extinction cross-section for a single AuNU with various tip numbers.

**Fig. S17** BEM simulation of the extinction cross-section for a single AuNU with various tip lengths.

**Fig. S18** BEM simulation of the extinction cross-section for a single AuNU with different core sizes.

**Fig. S19** BEM simulation of the plasmonic coupling properties for coupled AuNUs.

**Fig. S20** Influence of coupling angle between two anisotropic NPs on their optical properties.

**Fig. S21** Disposable spectrometer cuvettes (Fisher Scientific) made of clear plastic for smartphone testing.

Table S1 Antigen diagnostics for pathogens using plasmonic gold nanoparticles (NPs).

| Plasmonic NPs | Target | Detection method | Sample matrix | Amplification technique | LOD | Ref. |
| --- | --- | --- | --- | --- | --- | --- |
| Au nanosphere | Influenza virus | Colorimetric  SERS  LFA | Buffer  Buffer  Clinical | NA  Laser excitation  NA | 100 PFU  1.5 PFU  1000 PFU | ^1^ |
| Au nanosphere | Influenza virus H1N1/H3N2 | ELISA | Clinical | Magnetically assisted purification | 5×10^-4^ ng/mL for H1N1  2.5 PFU/mL for H3N2 | ^2^ |
| Au nanosphere | Hepatitis B surface antigen/α-fetoprotein | ELISA | Serum | Gold enhancement | 10^-3^ ng/mL | ^3^ |
| Hollow Au nanosphere | SARS-  CoV-2 | SERS | Buffer | Magnetically assisted purification/  Laser excitation | 3.4 PFU/mL | ^4^ |
| Au nanoplate | Influenza virus | SERS | Buffer | AuNP enhancement/  Laser excitation | 100 PFU/mL | ^5^ |
| Au nanorod | Hepatitis B virus antigen | ELISA | Buffer | Multiple washing steps | 0.01 IU/mL | ^6^ |
| Au nanobipyramids | Influenza virus H5N1 antigen | ELISA | Buffer/  Serum | Silver shell enhancement | 1 pg/mL | ^7^ |
| Au nanostar | Zika/Dengue virus antigen | LFA | Serum | SERS | 0.72 ng/mL for ZIKV  7.67 ng/mL for DENV | ^8^ |

**Table S2 Reagents for size-controlled synthesis of Au nanourchins and Au nanospheres.**

| Shape | NP size  (nm) | UP water  (mL) | HAuCl_4_·3H_2_O  2.5 E-02 M  (mL) | Na_3_CA·2H_2_O  1.5E-02 M  (mL) | 15 nm Seeds  2.23 nM  (mL) | Hydroquinone  2.5 E-02 M  (mL) |
| --- | --- | --- | --- | --- | --- | --- |
| Urchin | 65 | 88.610 | 1.000 | 0.499 | 0.911 | 12.000 |
|  | 75 | 88.665 | 1.000 | 0.499 | 0.456 | 12.000 |
| Sphere | 50 | 94.370 | 0.973 | 0.973 | 3.707 | 0.973 |
|  | 100 | 96.670 | 0.997 | 0.997 | 0.339 | 0.997 |

**Table S3 RSV A2 primer sequences for LAMP Assay.**^9^

| Primer name | Sequence (5′-3′) |
| --- | --- |
| FIP | TCTGCTGGCATGGATGATTGGAGACGATGATCCTGCATCA |
| BIP | CTAGTGAAACAAATATCCACACCCAGCACTGCACTTCTTGAGTT |
| LF | ACATGGGCACCCATATTGTAAG |
| LB | AGGGACCTTCATTAAGAGTCATGAT |
| F3 | GCTGTTCAATACAATGTCCTAGA |
| B3 | GGTAAATTTGCTGGGCATT |

Note: 10× primer mix was prepared with 16 μM FIP/BIP, 4 μM LF/LB, and 2 μM F3/B3.

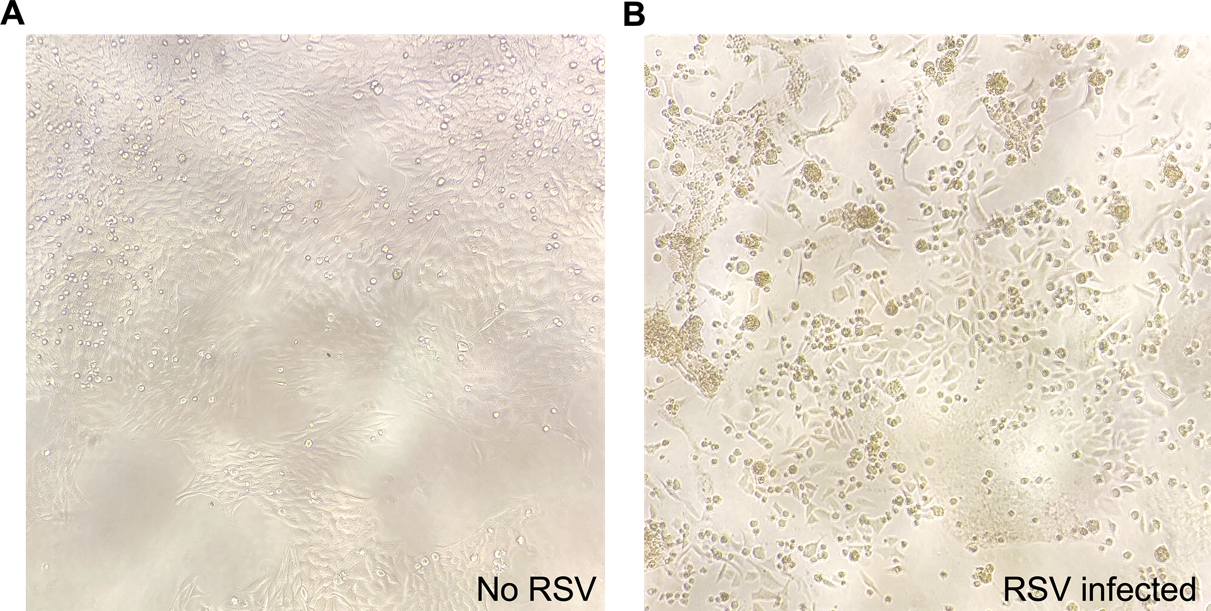

**Fig. S1 Large scale propagation of Respiratory syncytial virus (RSV) A2 stain.** Bright-field images of (A) HEp-2 cells without RSV infection, and (B) HEp-2 cells inoculated with RSV at day 6. Multinucleated cells (syncytia) are presented due to RSV-induced cell fusion.

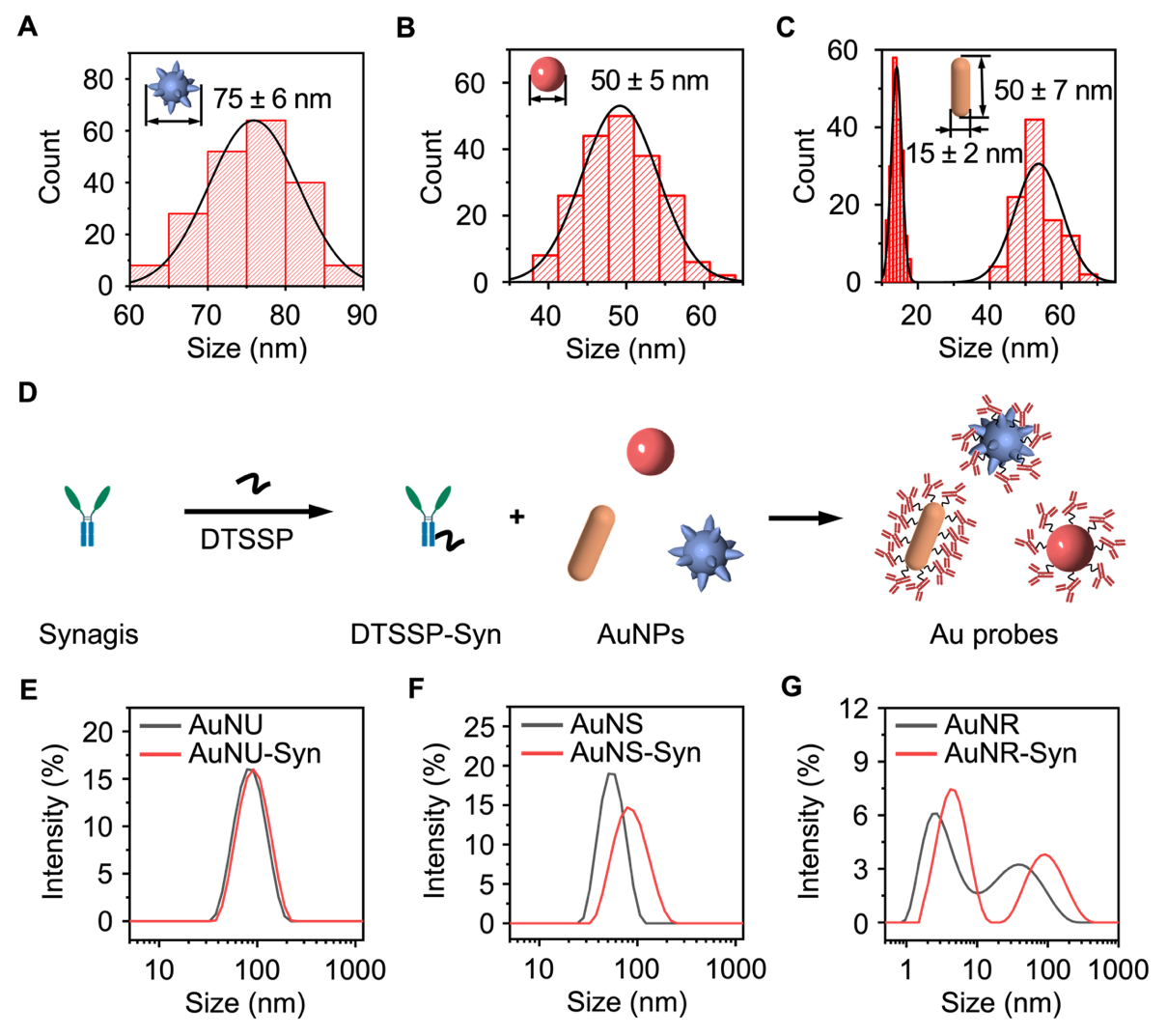

**Fig. S2 Characterization of AuNP probes.** Size distribution profile of (A) Au nanourchins (AuNUs), (B) Au nanospheres (AuNSs) and (C) Au nanorods (AuNRs) by randomly measuring 200 particles in TEM images. The average particle dimensions were measured as 75 nm ± 6 nm for AuNUs (defined as tip-to-tip length), 50 nm ± 5 nm for AuNSs, and 50 nm ± 7 nm and 15 nm ± 2 nm for the length and width of AuNRs. (D) Schematic illustration of bioconjugation of Synagis to AuNP probes via a grafting-to approach. Dynamic light scattering (DLS) measurements of (E) AuNUs, (F) AuNSs, and (G) AuNRs before and after conjugation with Synagis.

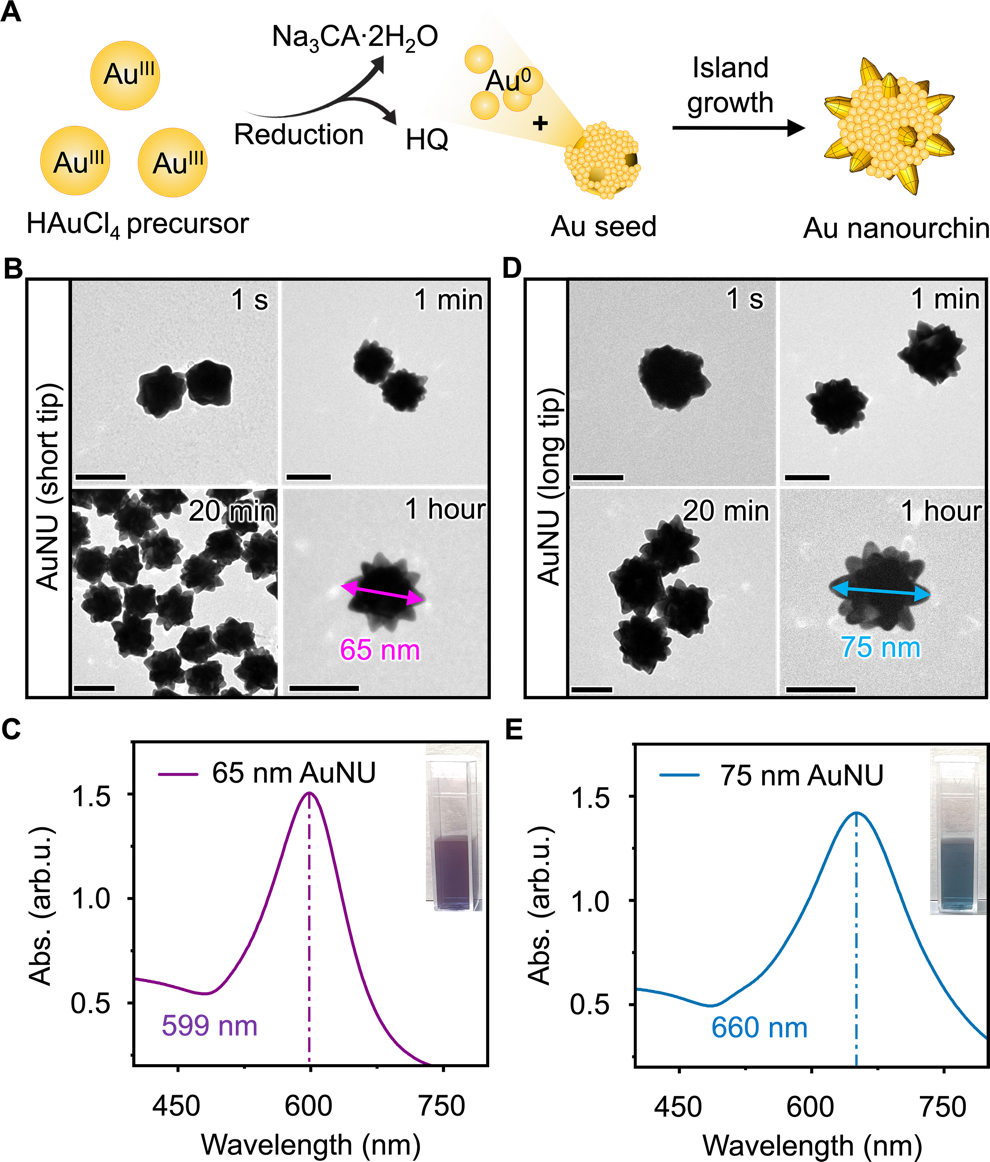

**Fig. S3 Synthesis and characterization of AuNUs of different sizes.** (A) Schematic illustration of the seed-mediated growth of Au nanourchins. (B, D) Transmission electron microscopy images of (B) 65nm AuNU with short tips and (D) 75 nm AuNU with long tips. (C, E) Absorbance spectra were measured for (C) 65 nm AuNU and (E) 75 nm AuNU with. Insets in C and E show the photographs of the corresponding nanoparticle solutions.

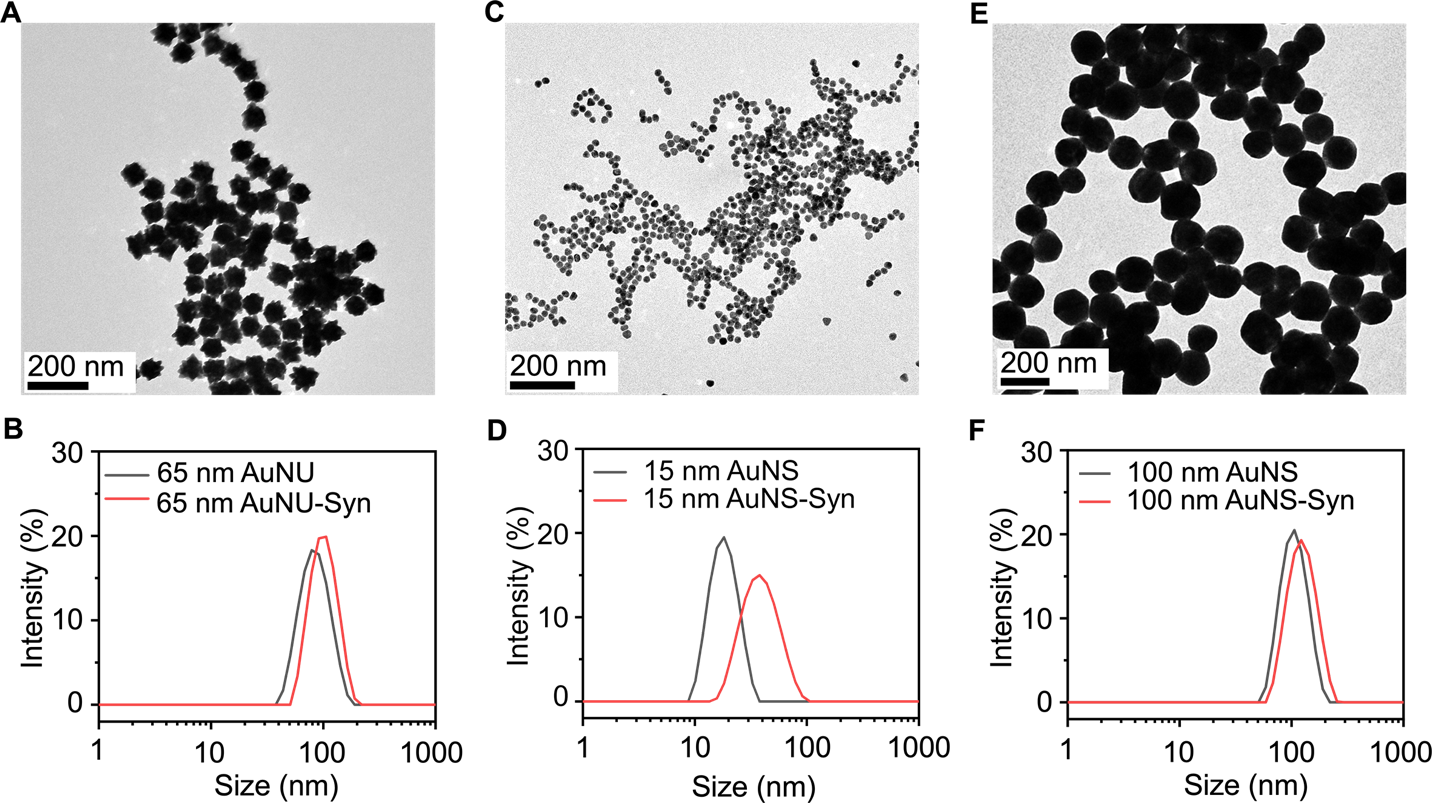

**Fig. S4 Characterization of 65 nm AuNUs with short tips and different sizes of AuNSs.** (A, C, E) Transmission electron microscopy images of (A) 65 nm AuNUs, (C) 15 nm AuNSs and (E) 100 nm AuNSs. (B, D, F) Dynamic light scattering (DLS) of (B) 65 nm AuNUs, (D) 15 nm AuNSs and (F) 100 nm AuNSs before and after conjugation with Synagis.

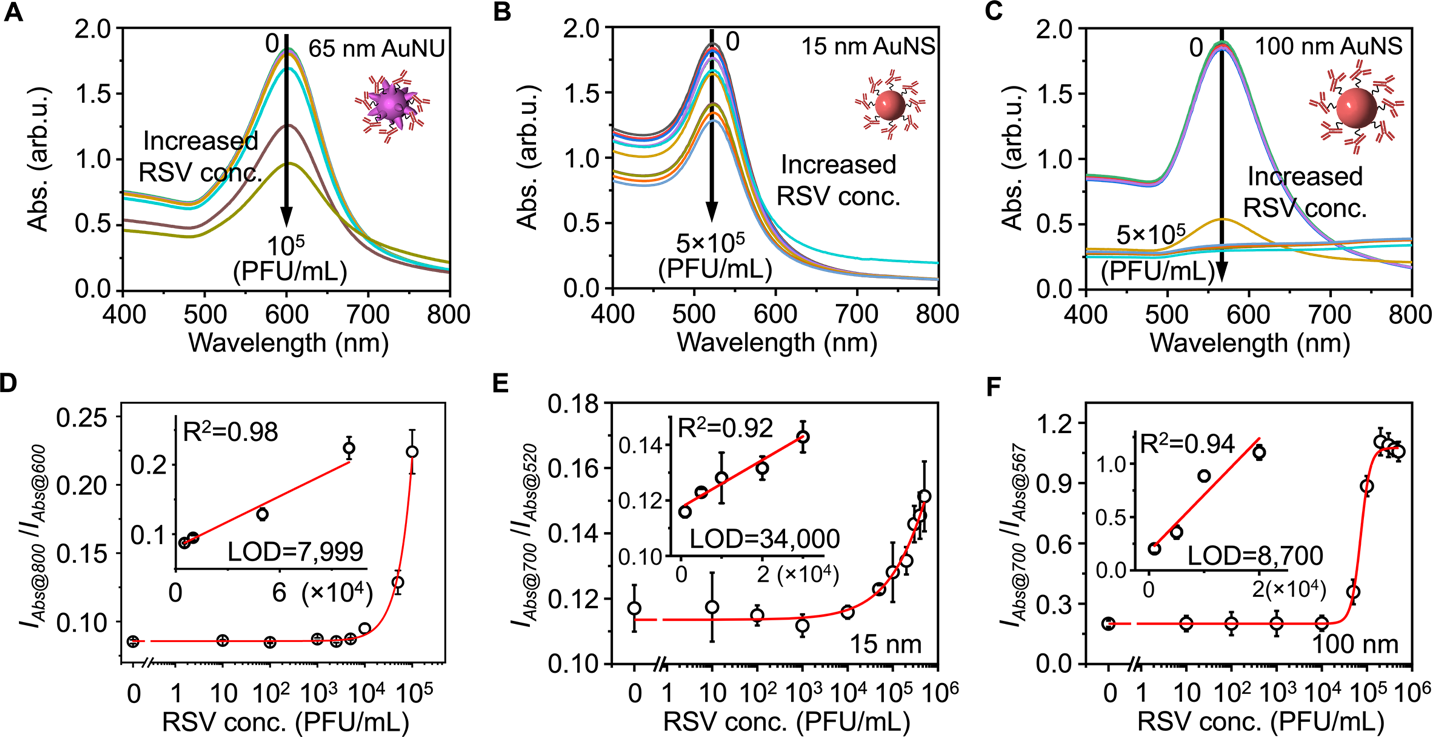

**Fig. S5 Respiratory syncytial virus (RSV) detection using plasmonic AuNUs and AuNSs with different sizes.** (A-C) Colorimetric detection using (A) 65 nm AuNUs, (B) 15 nm and (C) 100 nm AuNS probes, respectively. Each inset shows the model of the probe for detection of RSV with serial dilutions (0-100,000 PFU/mL for AuNUs, 0-500,000 PFU/mL for AuNSs). (D-F) Corresponding calibration curves by plotting the absorbance intensity (*I_Abs_*) ratio at two wavelengths against RSV titers. The error bars indicate the standard deviations (*n* = 3). Insets in (D-F) show the linear detection range and calculated limit of detection (LOD).

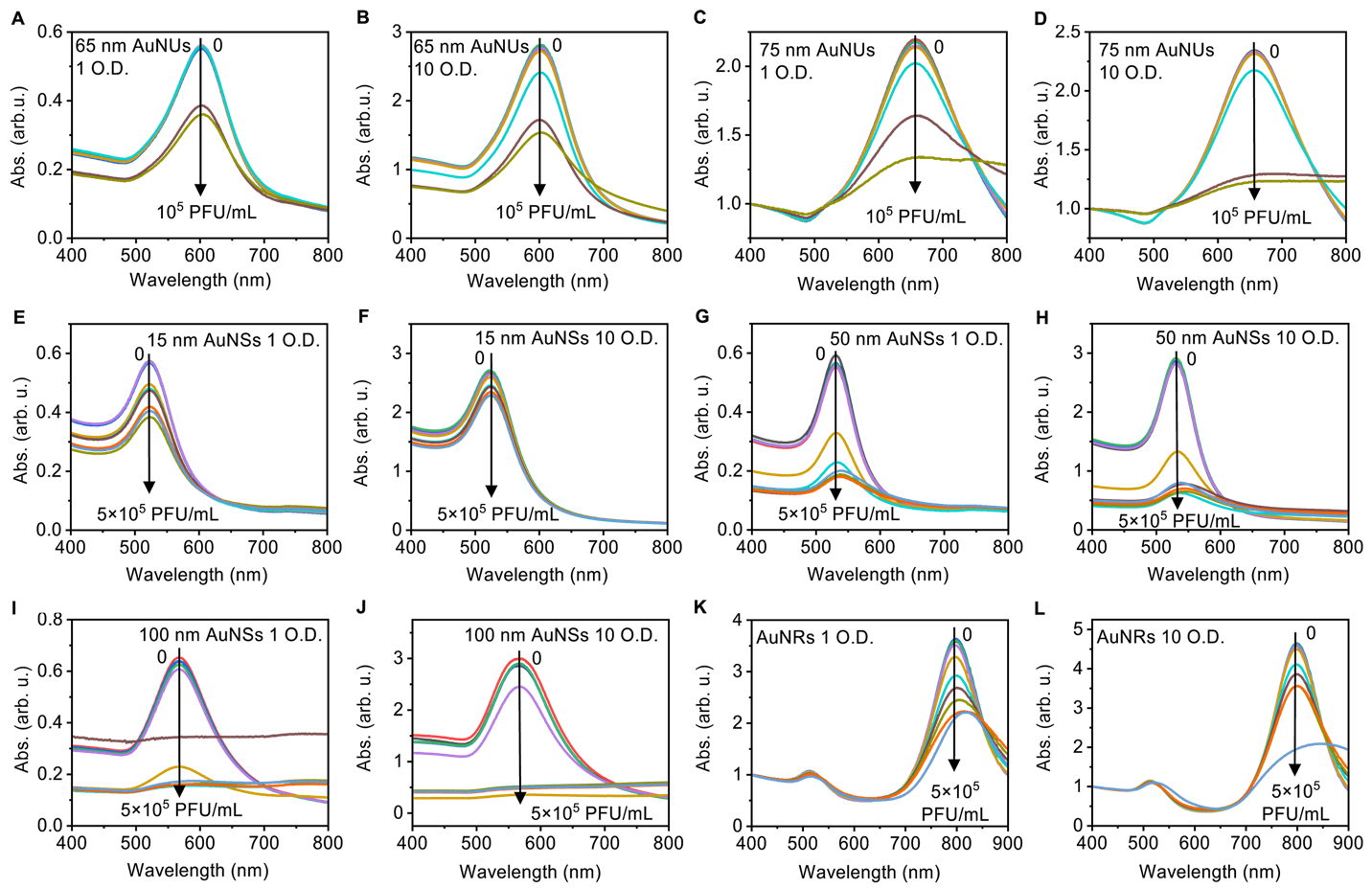

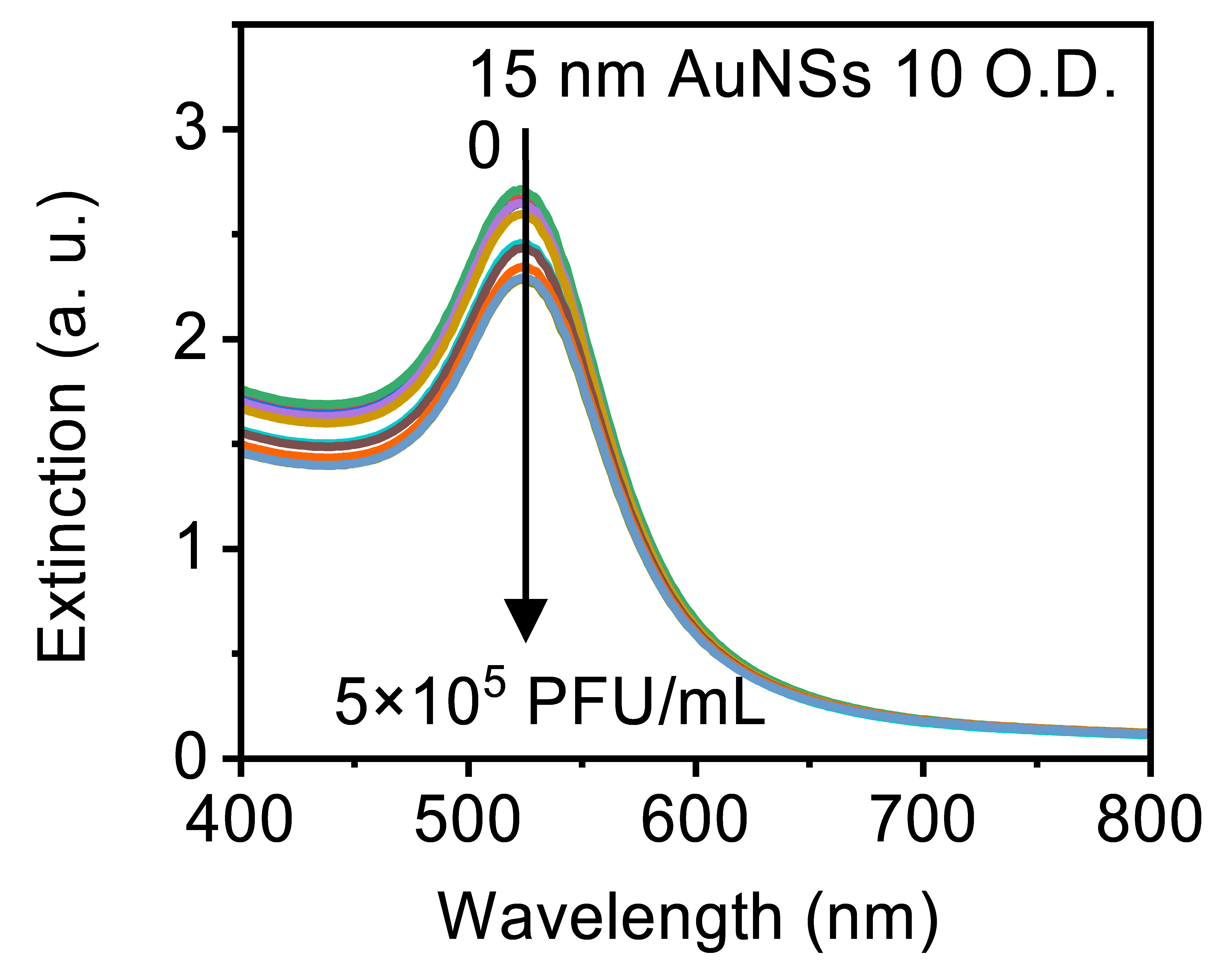

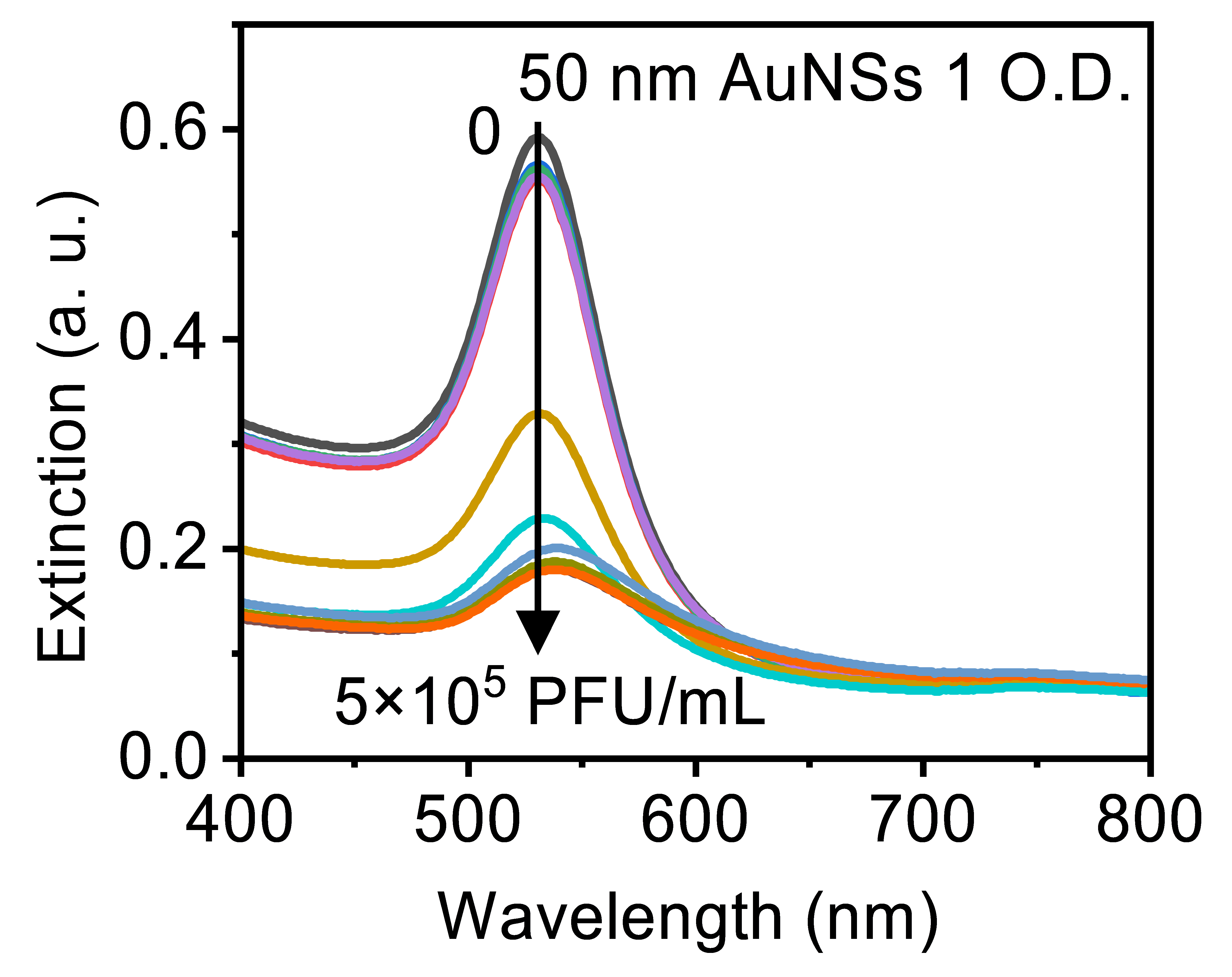

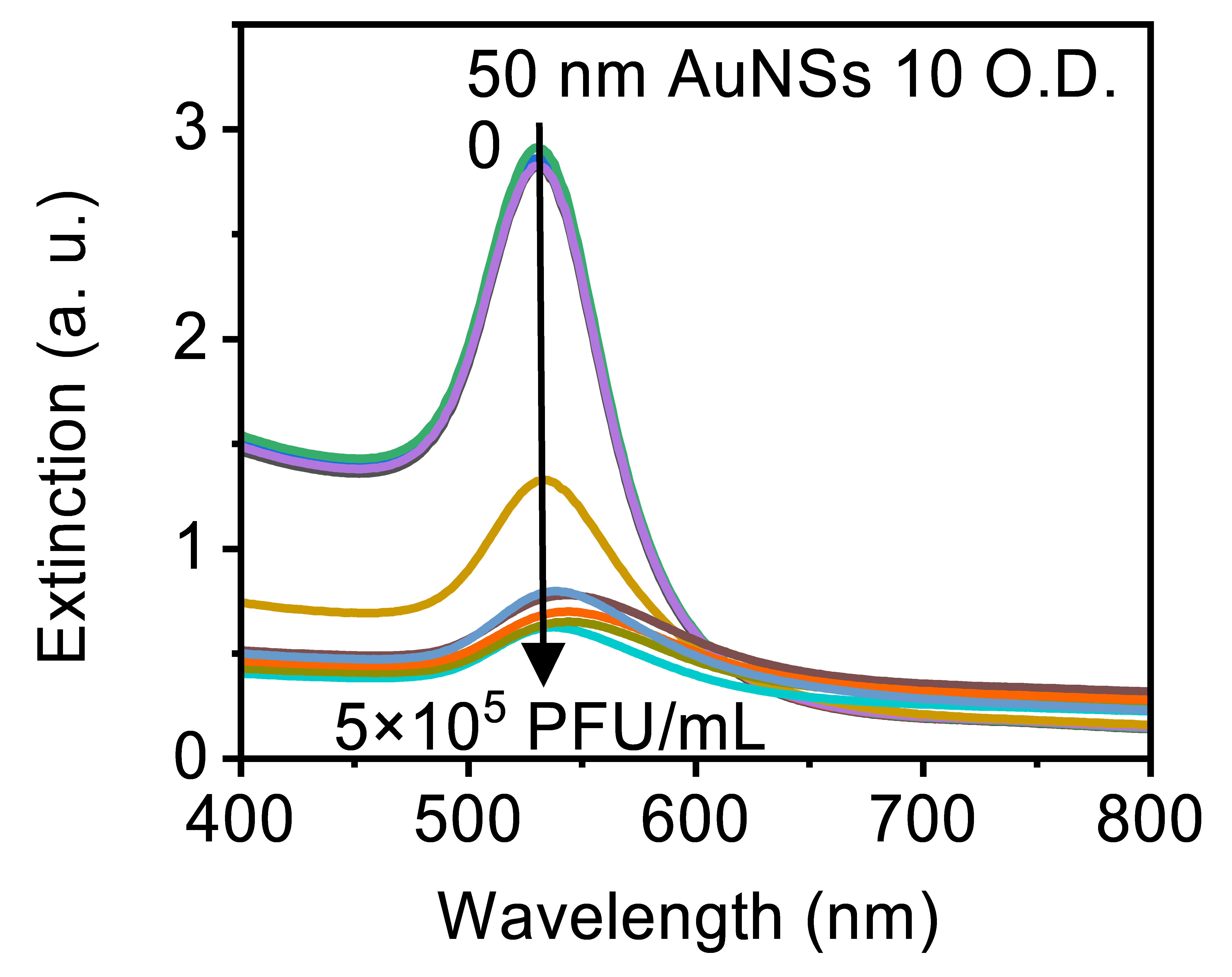

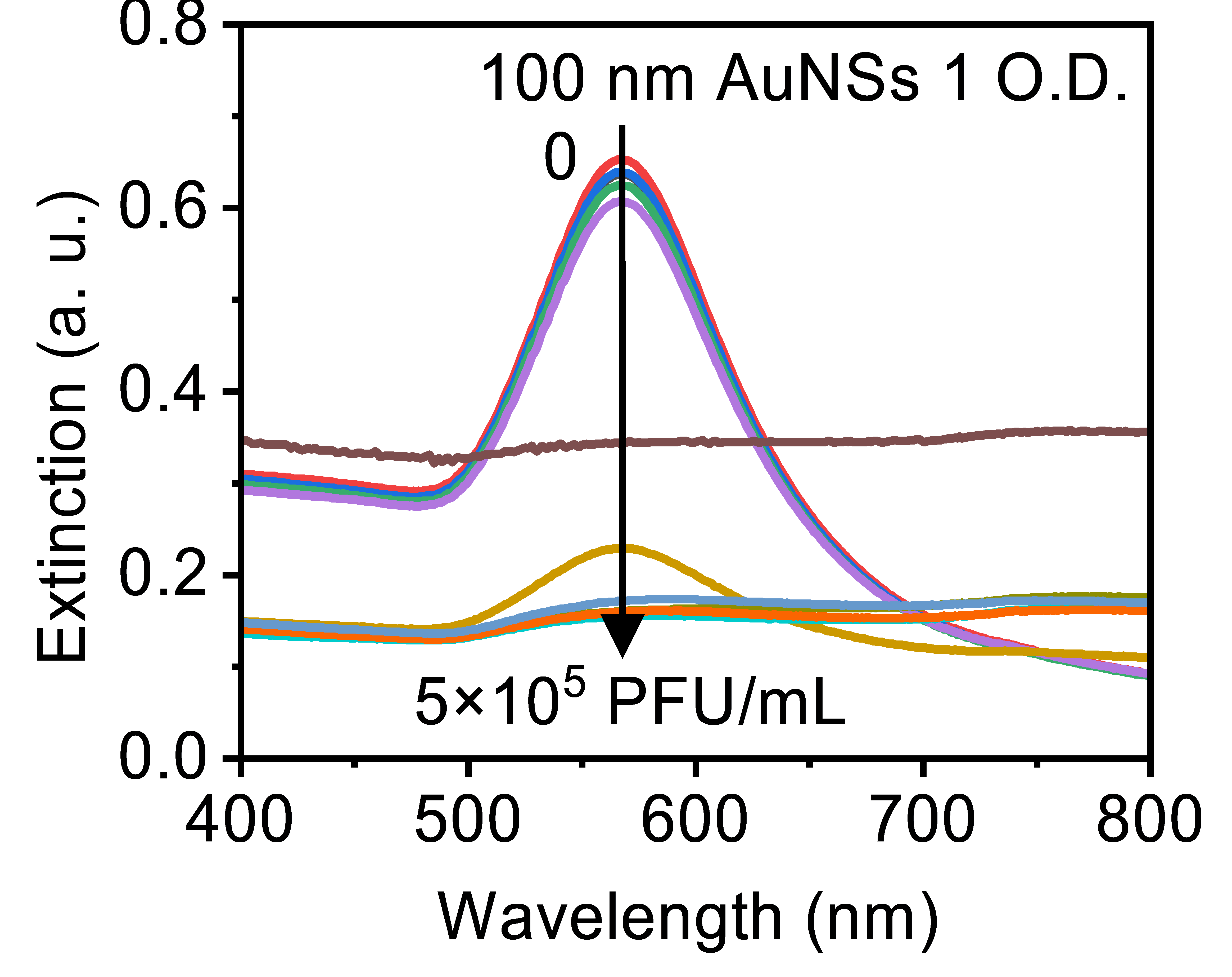

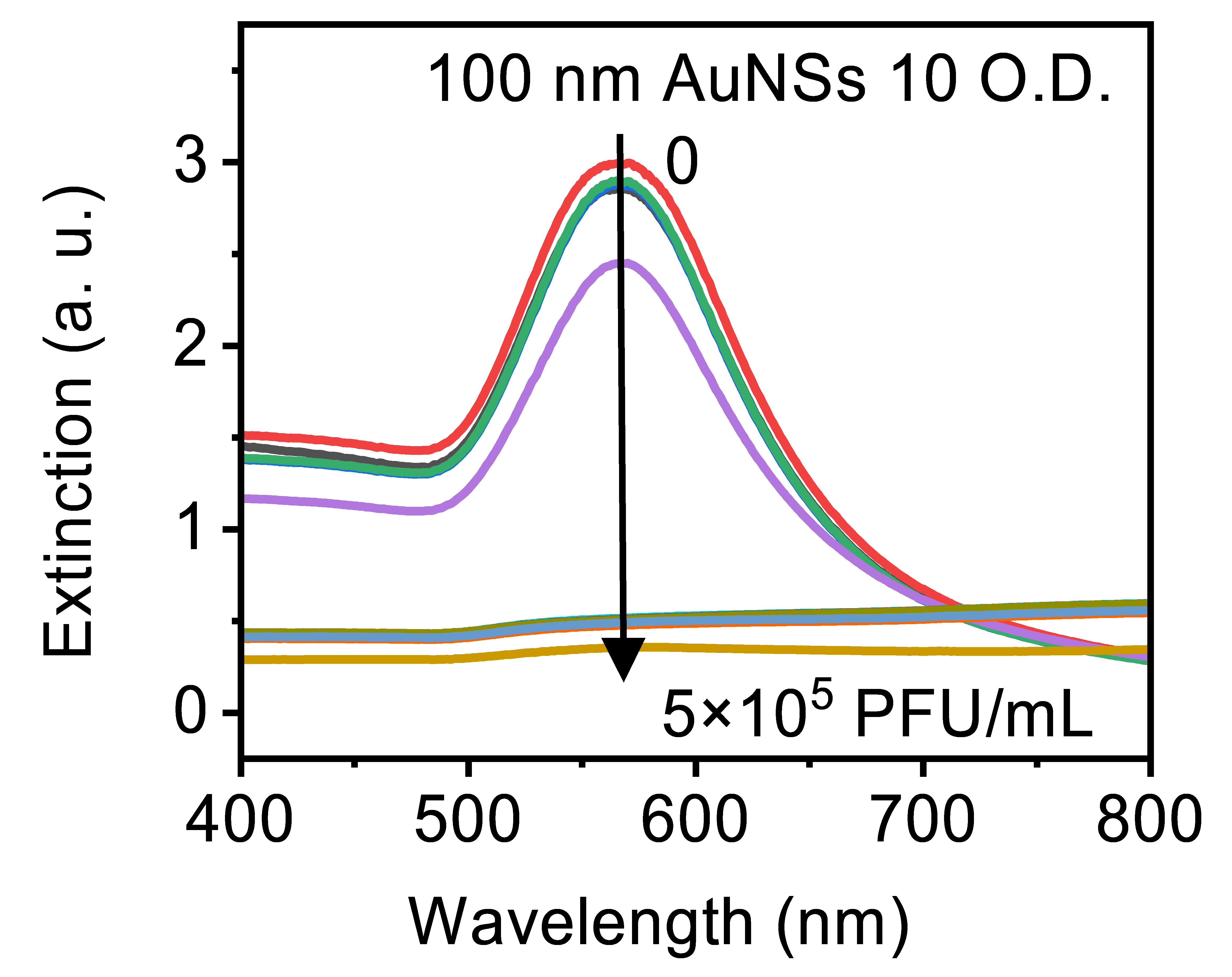

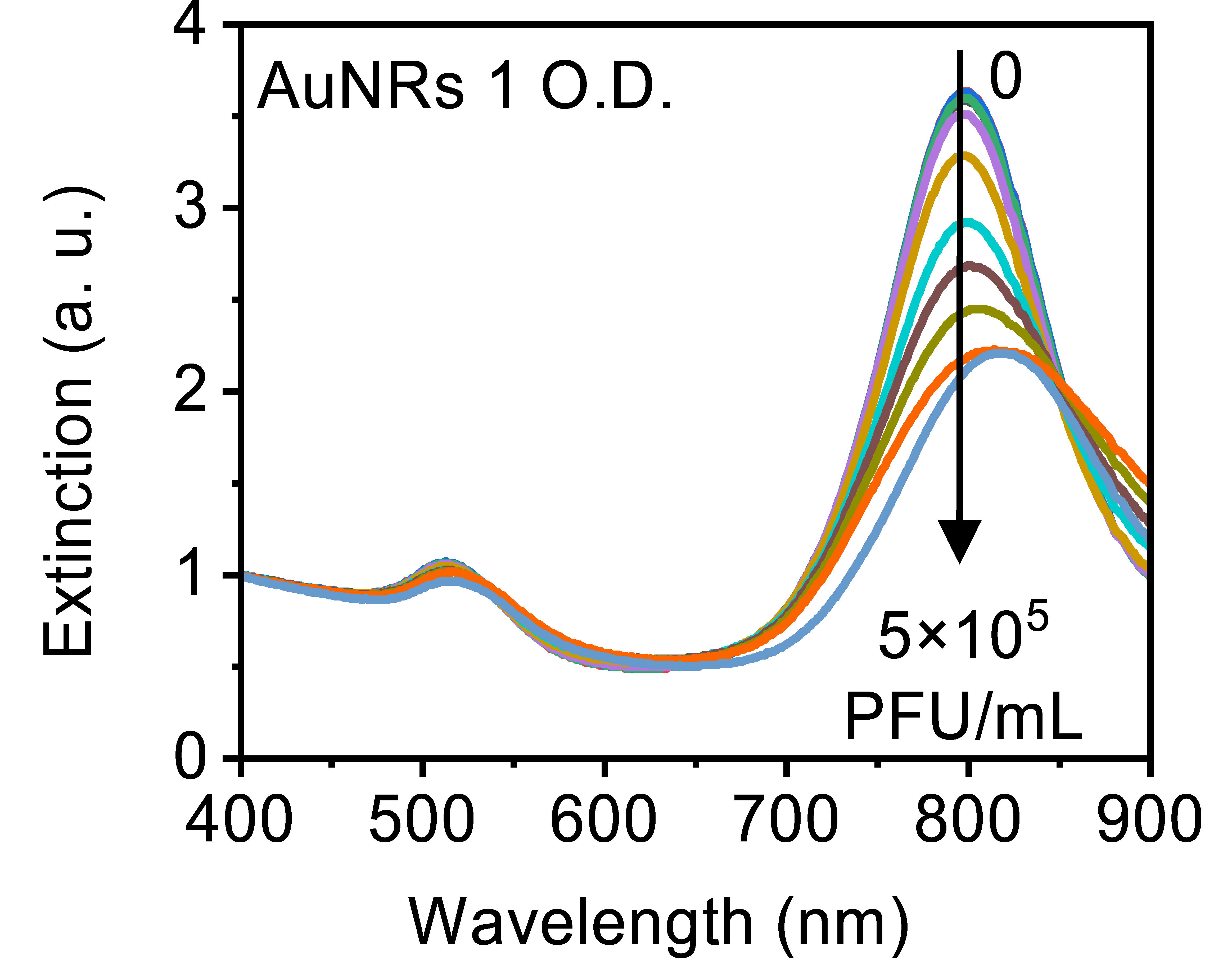

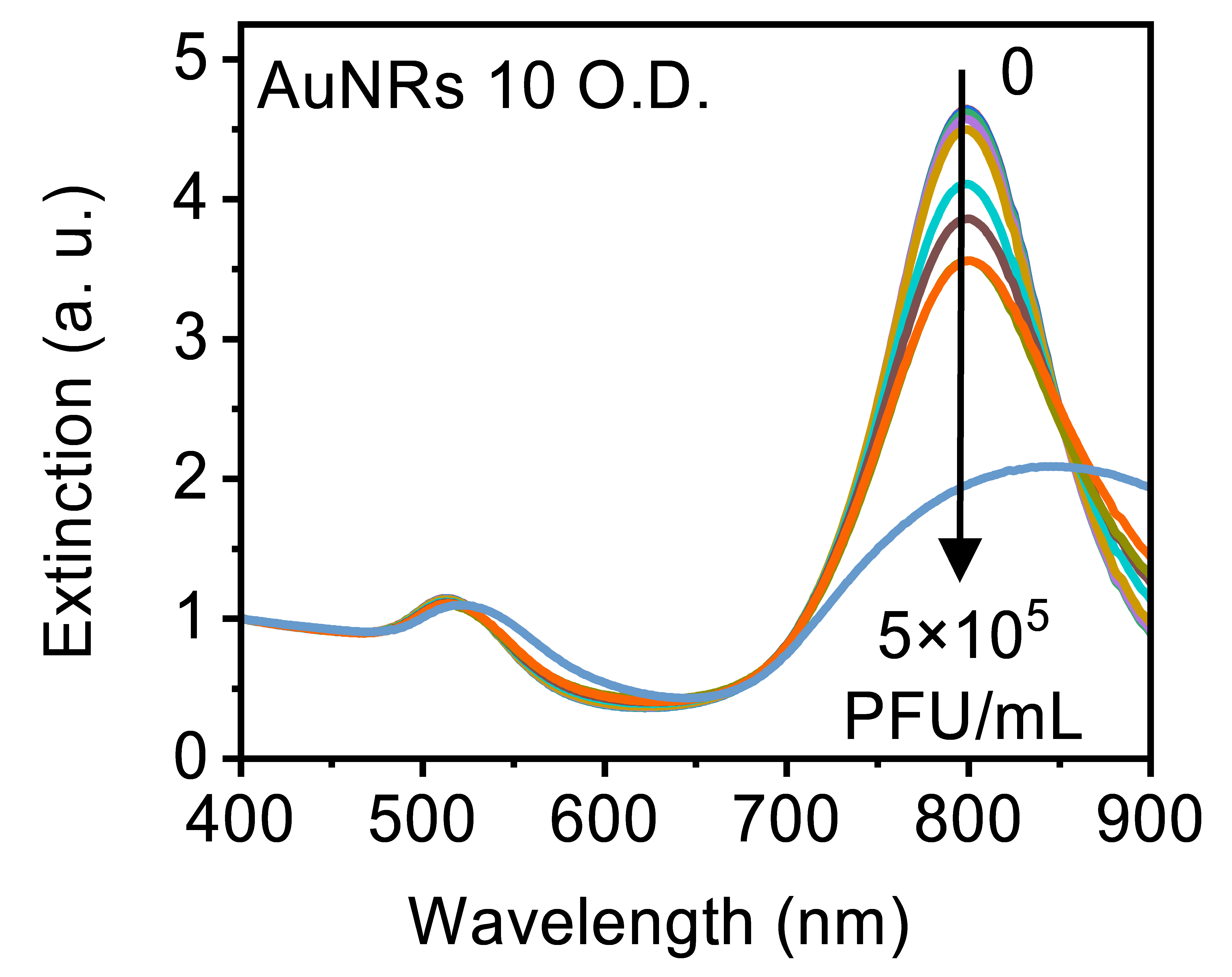

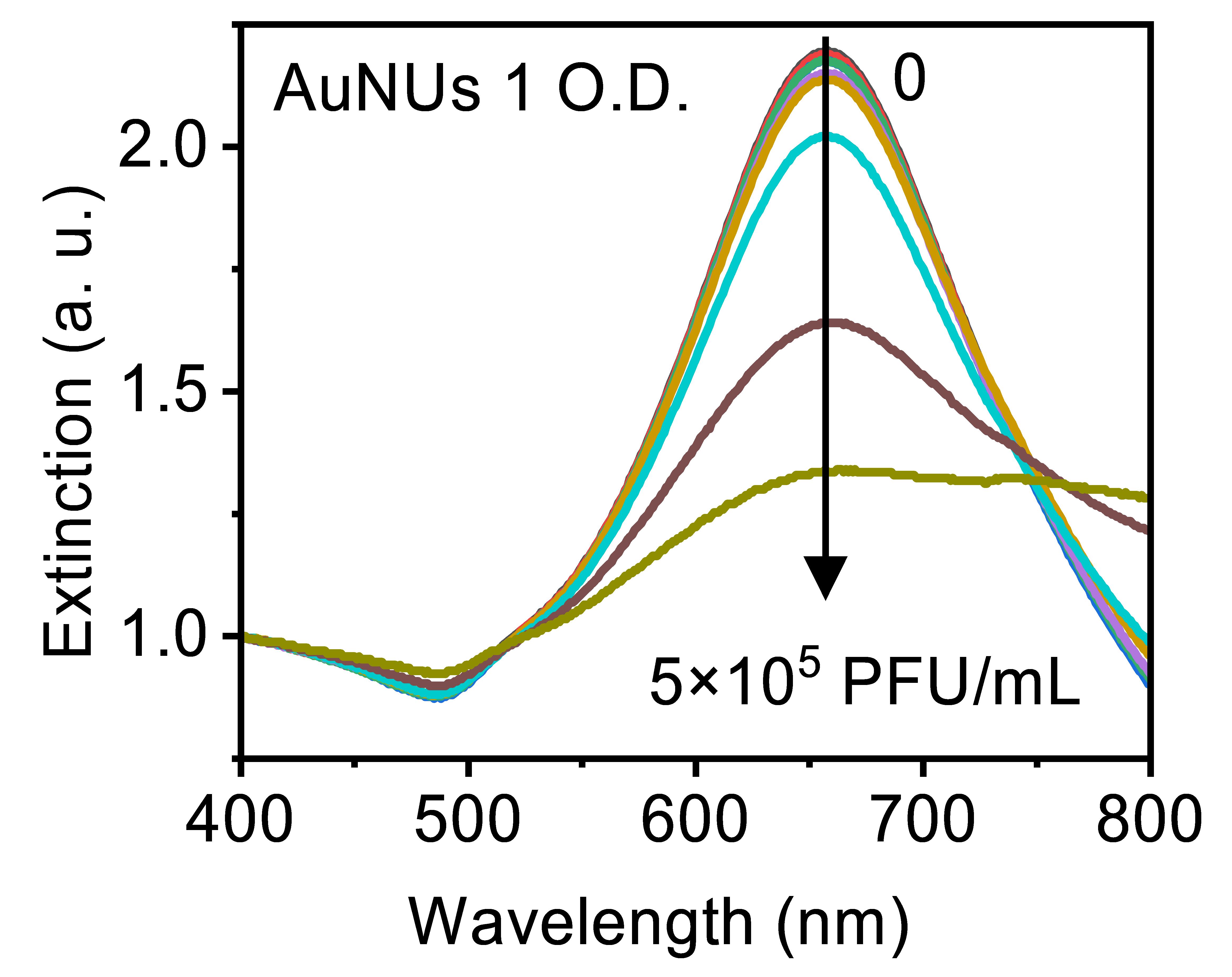

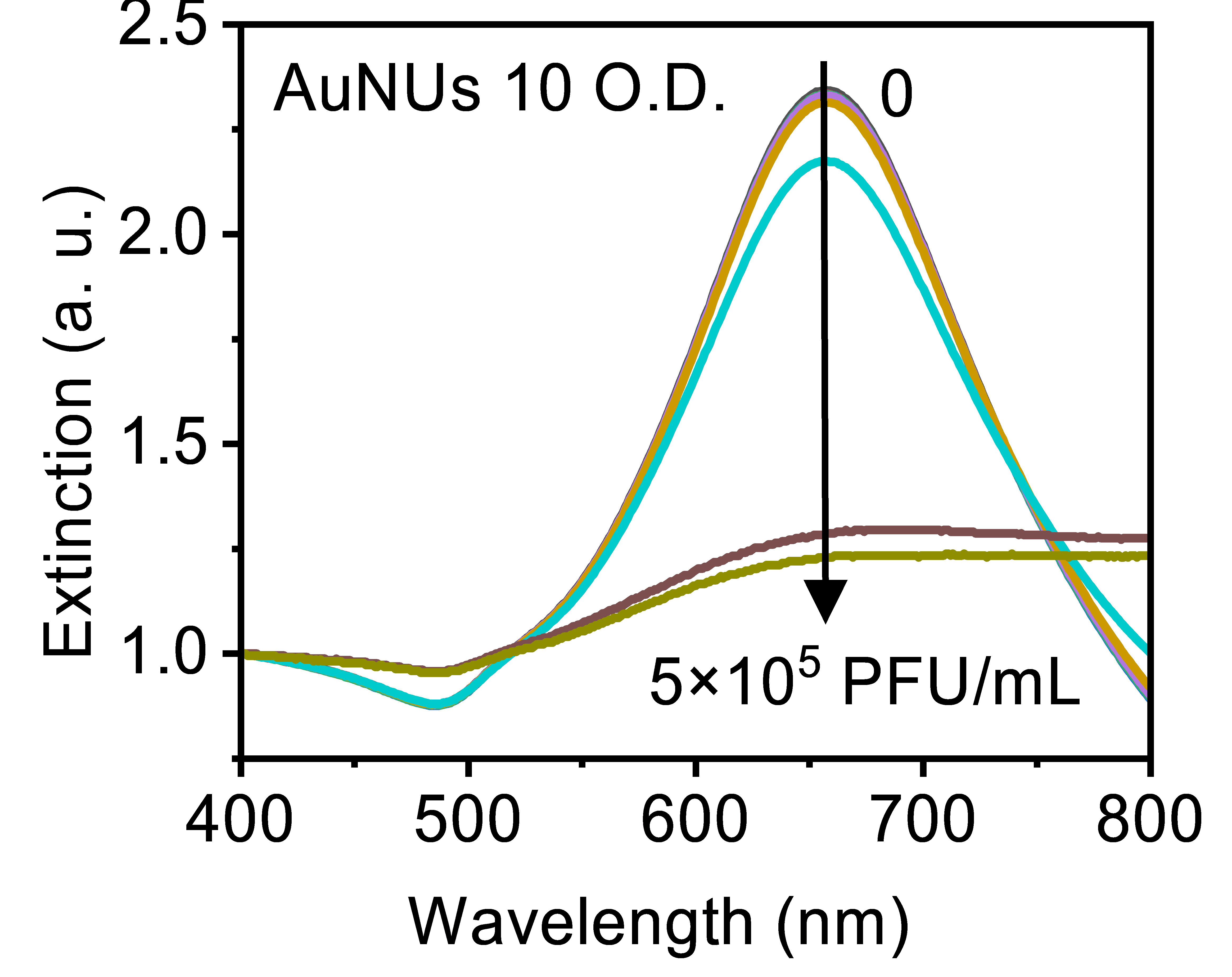

**b**

**Fig. S6 Respiratory syncytial virus (RSV) detection using AuNP probes of different concentration.** (A-L) Spectra measurements for 1 O.D. and 10 O.D. of AuNPs incubated with a serial dilution of RSV.

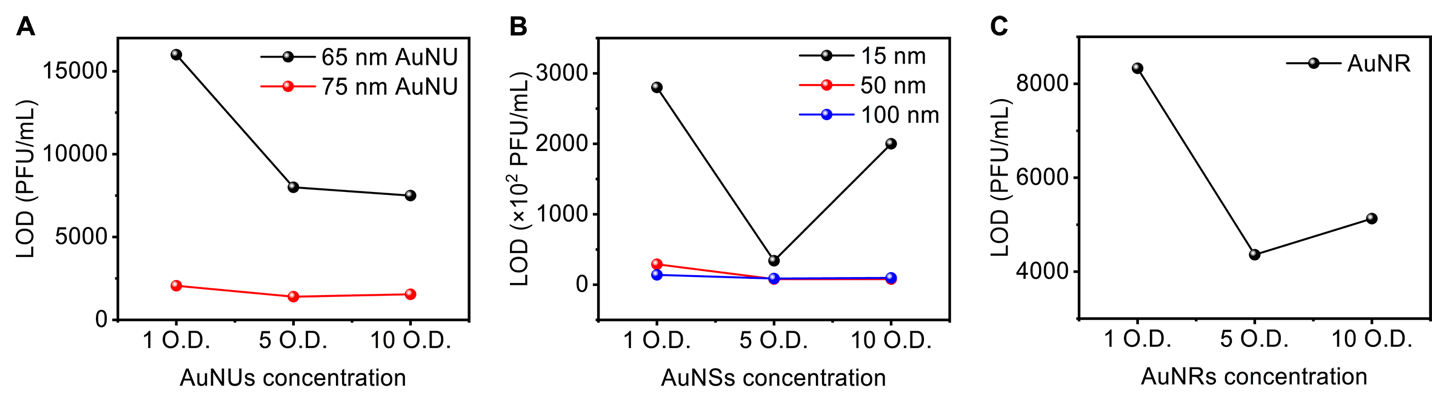

**Fig. S****7 Effect of AuNP probes concentration on RSV detection.** O.D.= Optical density. Limit of detection (LOD) for RSV by (A) AuNUs, (B) AuNSs, and (C) AuNRs. Higher O.D. of AuNP probes offer lower LOD, but the trend was saturated at 5 O.D.

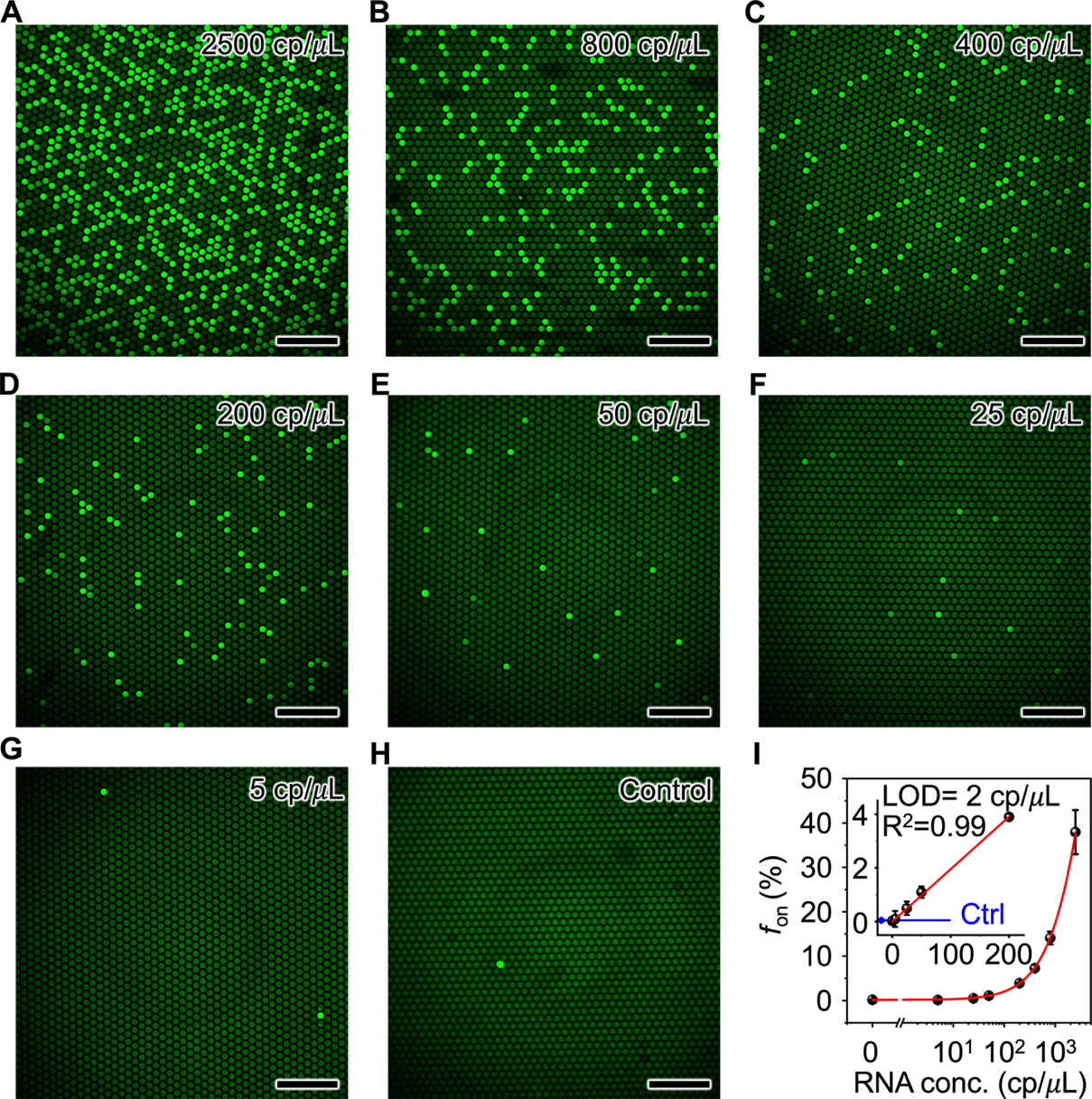

**Fig. S8 dLAMP of quantitative genomic RNA for RSV A2 strain.** (A-H) Fluorescent images of on-chip LAMP reactions for serial dilutions of RNA, where ultra-pure water serves as the negative control. Scale bar = 500 um. (I) Positive “ON” signal (*f*_on_) counting results as a function of RNA concentration (copies per microliter). The error bars indicate the standard deviations (*n* = 3). Images were analyzed via MATLAB script. Inset shows the limit of detection (LOD) of the quantitative RNA for RSV A2.

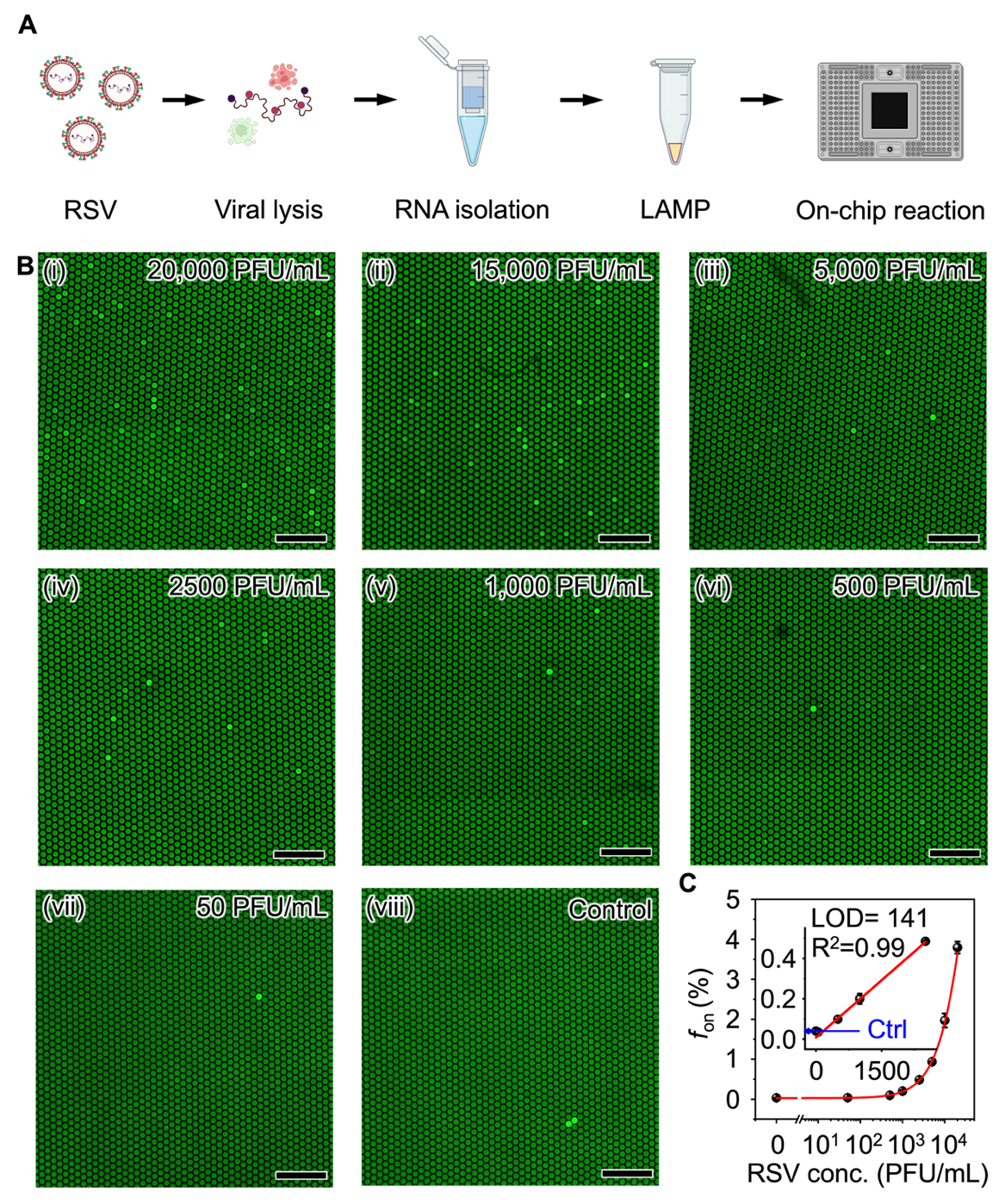

**Fig. S9 dLAMP assay for detection of RNA extracts from RSV A2 strain.** (A) Schematic of the complete assay process. (B) Fluorescent images of on-chip LAMP reactions for serial dilutions of RNA extracts from RSV A2, where ultra-pure water serves as the negative control. Scale bar = 500 um. (C) Positive “ON” signal (*f*_on_) counting results as a function of RSV concentration (Plaque-forming units per milliliter). The error bars indicate the standard deviations (*n* = 3). Images were analyzed via MATLAB script. Inset shows the limit of detection (LOD) for RSV A2.

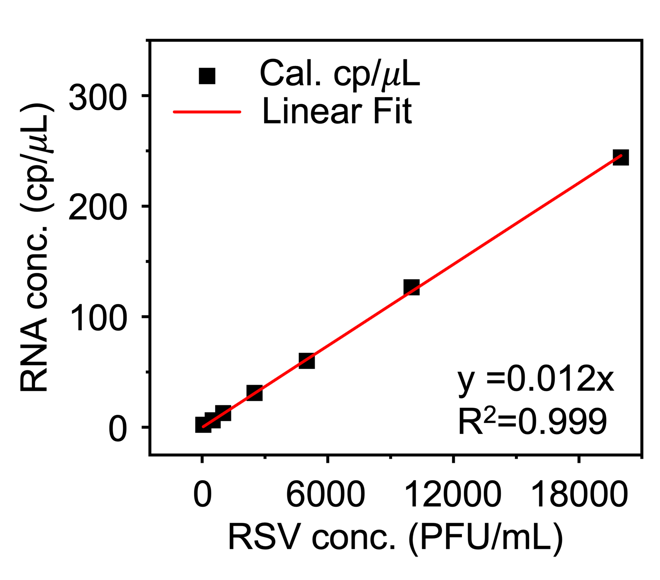

**Fig. S10 Calibration of RSV A2 and corresponding RNA concentration by dLAMP assay.** Based on *f*_on_ counting results for quantitative RNA (**Fig. S8**) and RNA extracts from RSV A2 (**Fig. S9**), we estimated 1 PFU/mL of RSV A2 is equal to approximately 0.012 copies/µL of RNA.

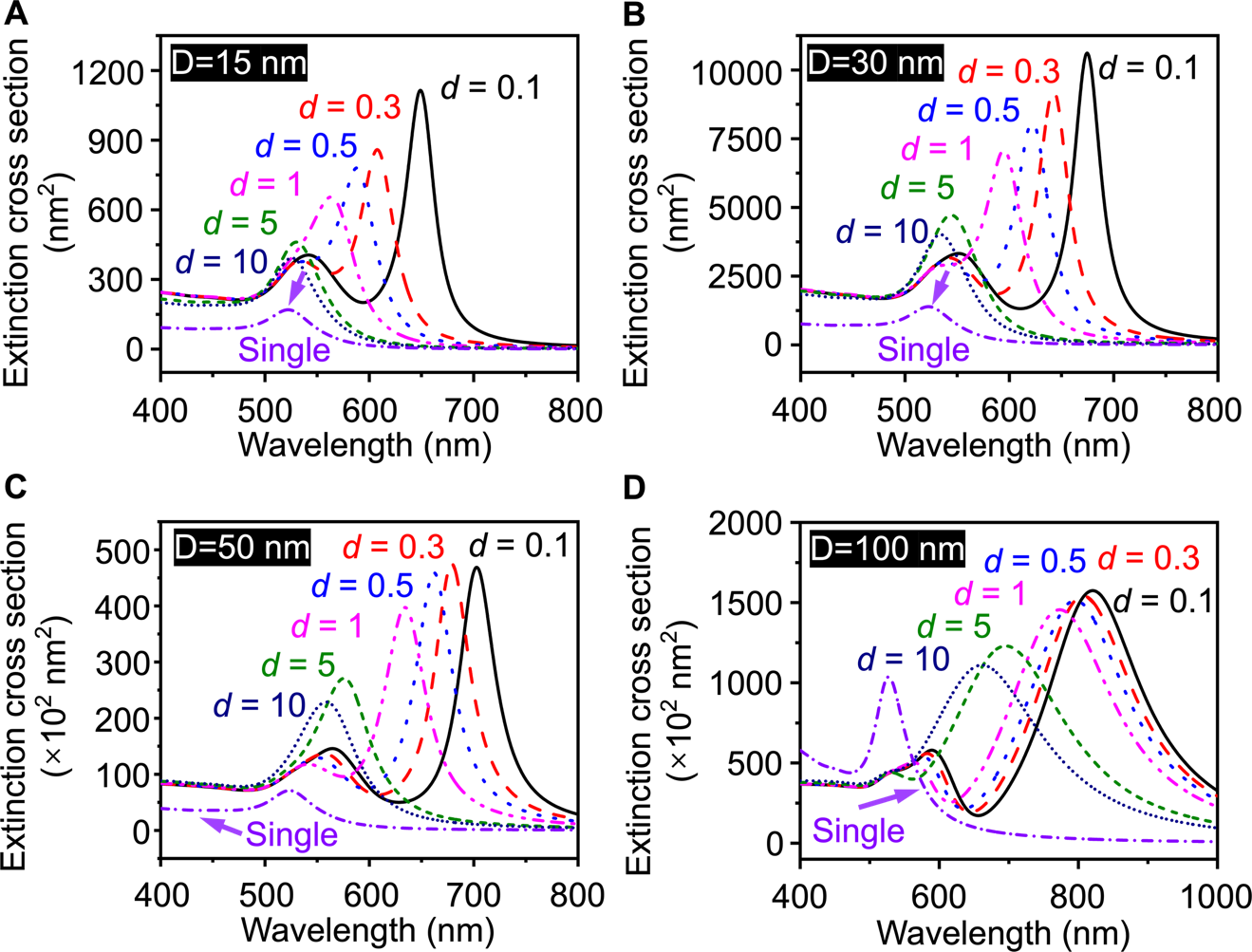

**Fig. S11 BEM simulation of the plasmonic coupling properties for AuNSs.** Simulated extinction cross-section at different interparticle distance for AuNSs with size of (A) 15 nm, (B) 30 nm, (C) 50 nm, and (D) 100 nm.

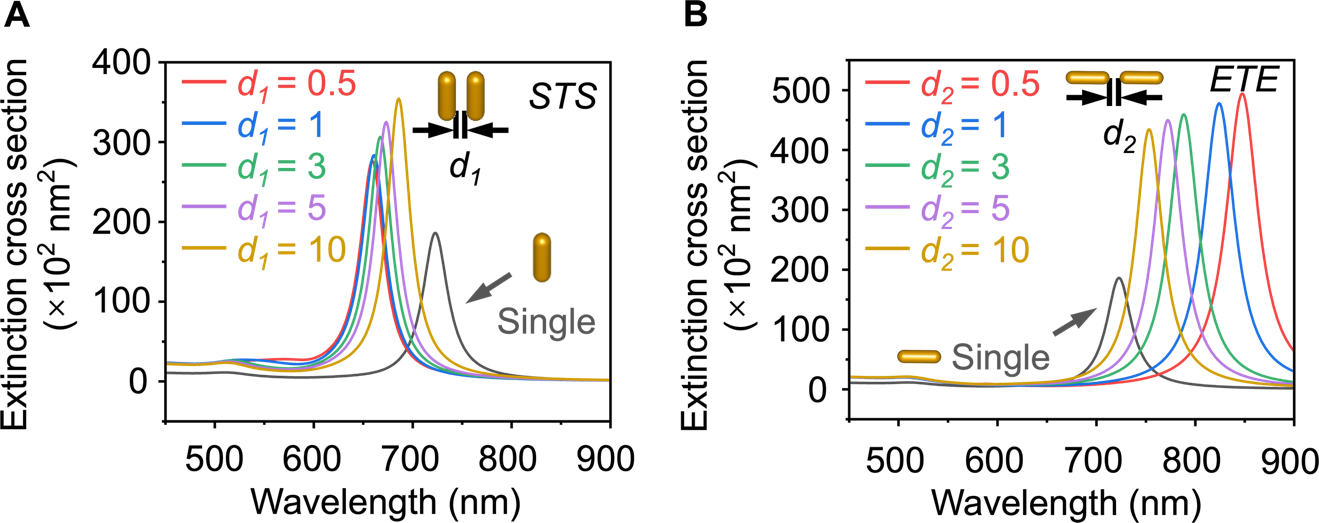

**Fig. S12 BEM simulation of the plasmonic coupling properties for AuNRs.** Simulated extinction cross-section at different interparticle distance for AuNRs with coupling orientation of (A) side-to-side (STS) and (B) end-to-end (ETE).

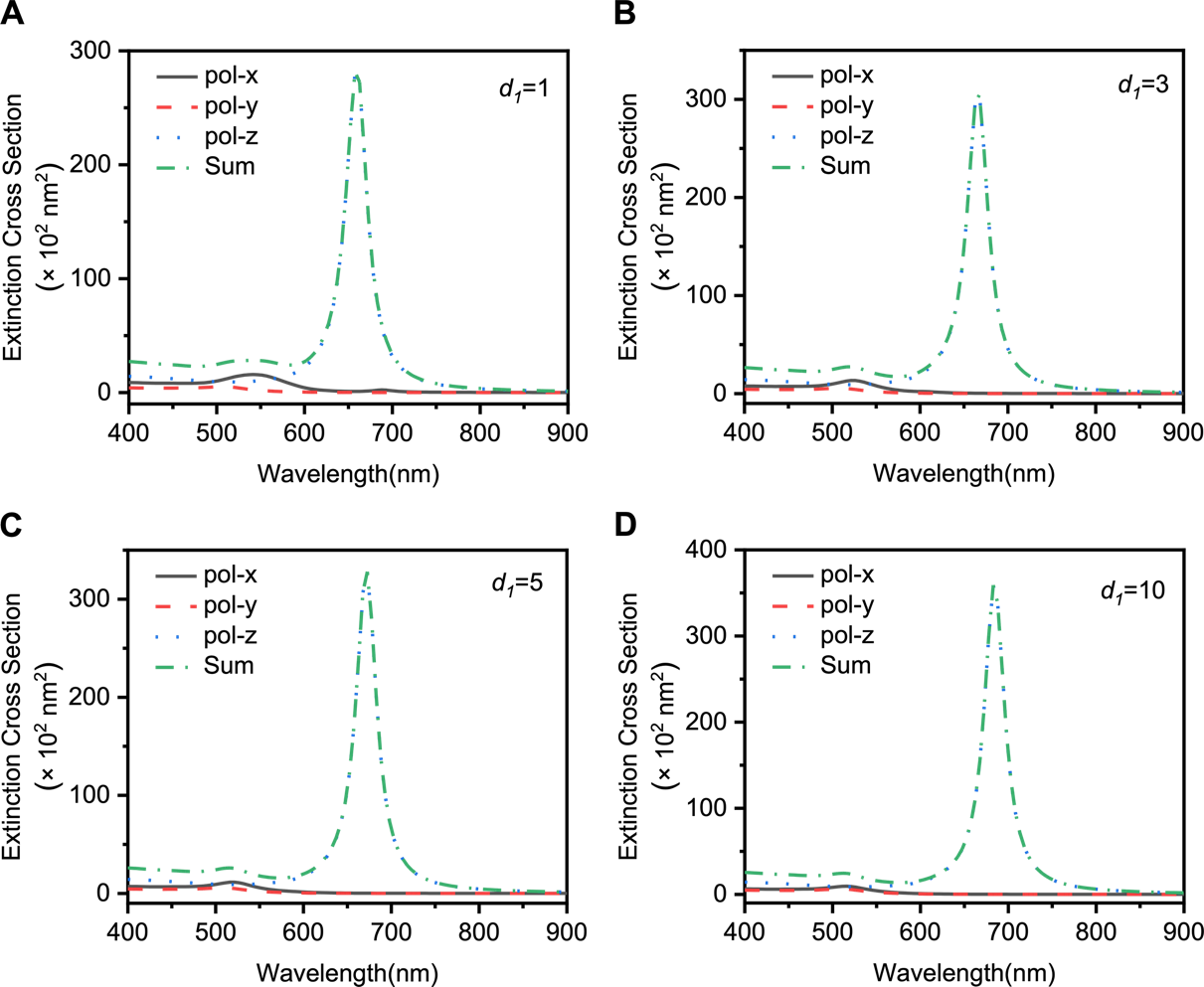

**Fig. S13 BEM simulation of the extinction cross-section for STS-coupled AuNRs with x-, y-, and z- polarization directions.** The interparticle distance *d_1_* was altered from (A) *d_1_* =1, (B) *d_1_* =3, (C) *d_1_* =5, to (D) *d_1_* =10 nm. The summation of extinction cross-section from three linear polarization directions are calculated for each case.

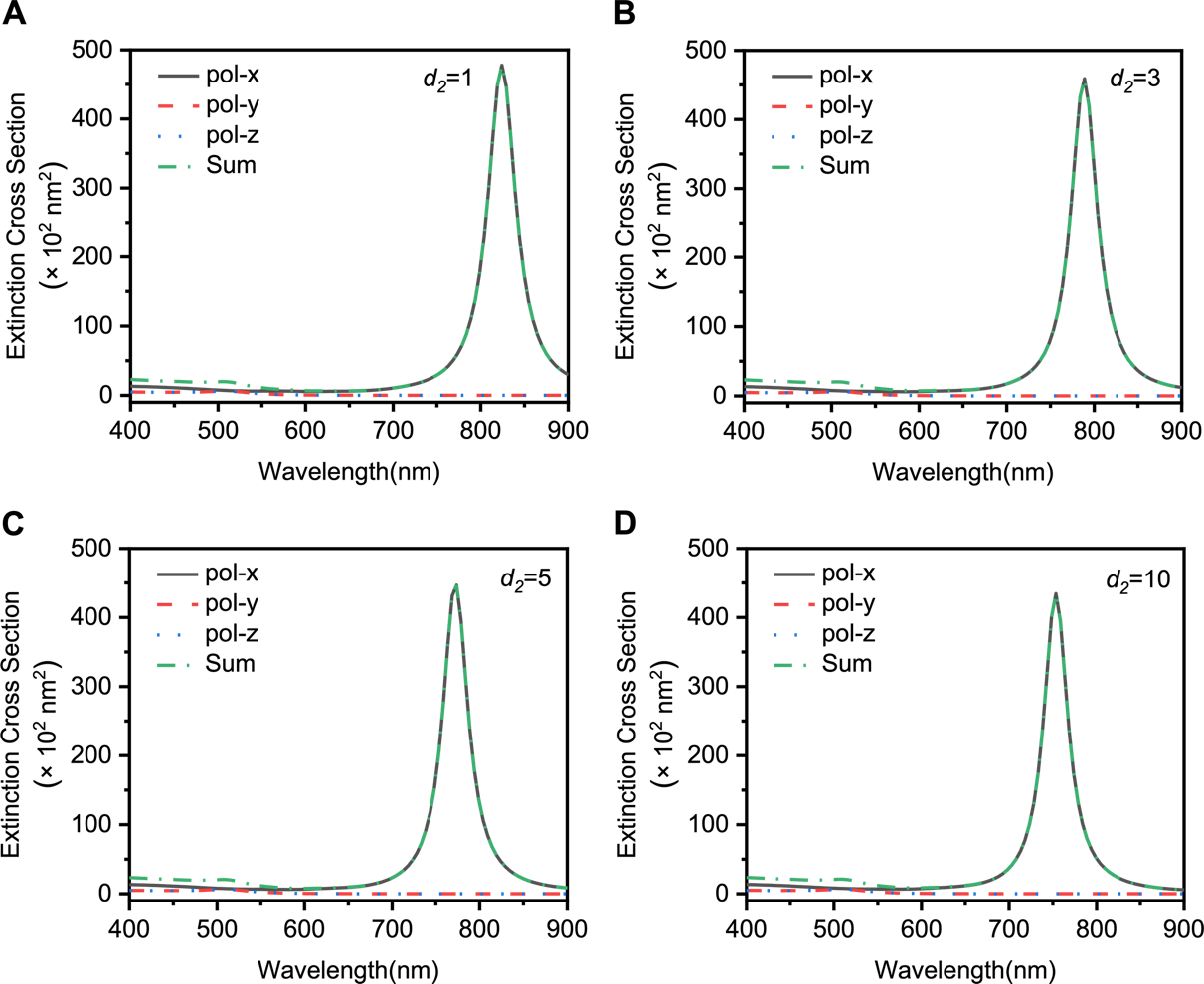

**Fig. S14 BEM simulation of the extinction cross-section for ETE-coupled AuNRs with x-, y-, and z- polarization directions.** The interparticle distance *d_2_* was altered from (A) *d_2_* =1, (B) *d_2_* =3, (C) *d_2_* =5, to (D) *d_2_* =10 nm. The summation of extinction cross-section from three linear polarization directions is calculated for each case.

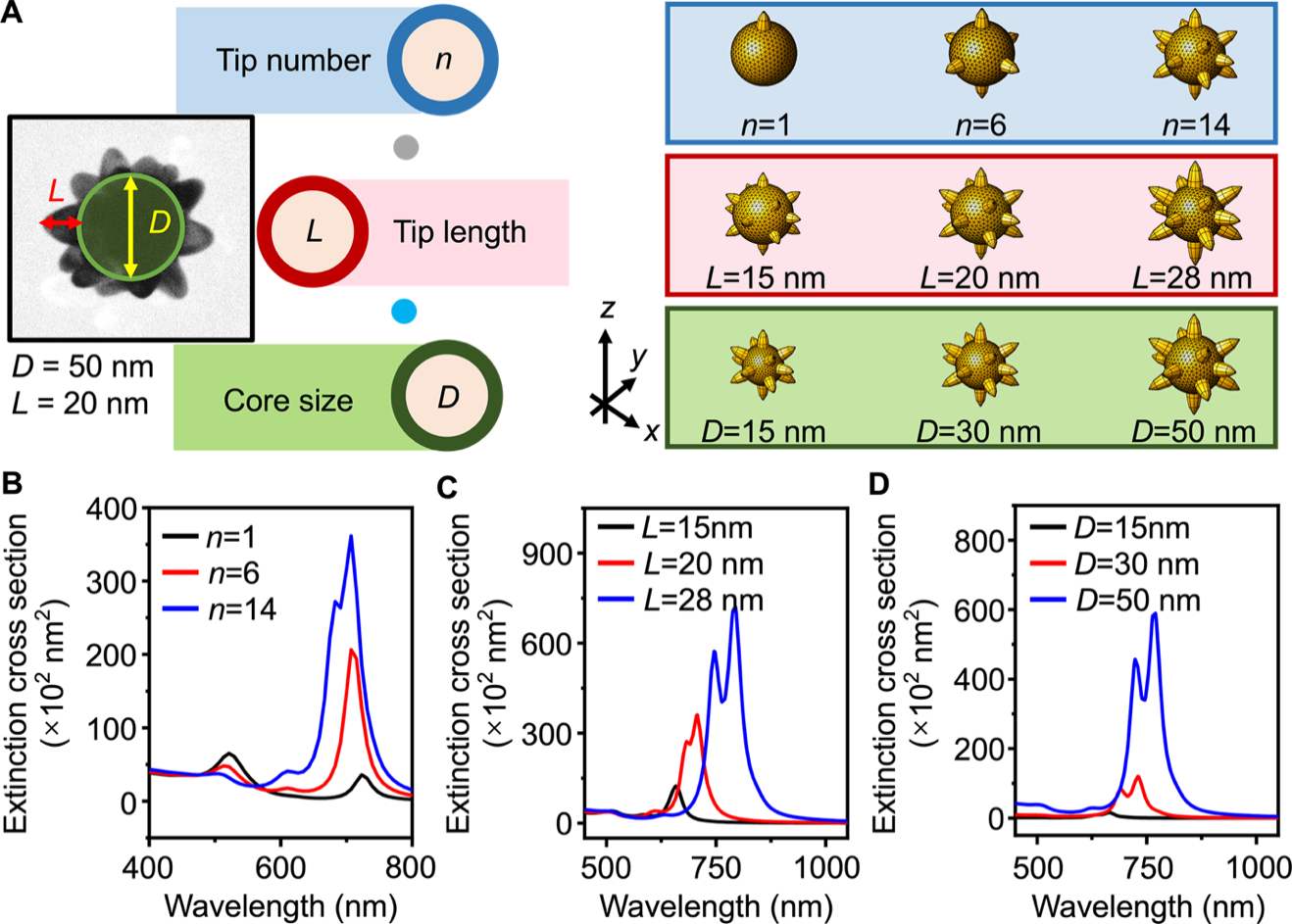

**Fig. S15 BEM simulation of plasmonic properties of AuNUs.** (A) Models for a single AuNU obtained from a TEM image with variable parameters, including tip number (*n*), tip length (*L*), and core size (*D*). (B-D) Simulated extinction cross-section for a single AuNU with varied *n*, *L* and *D*.

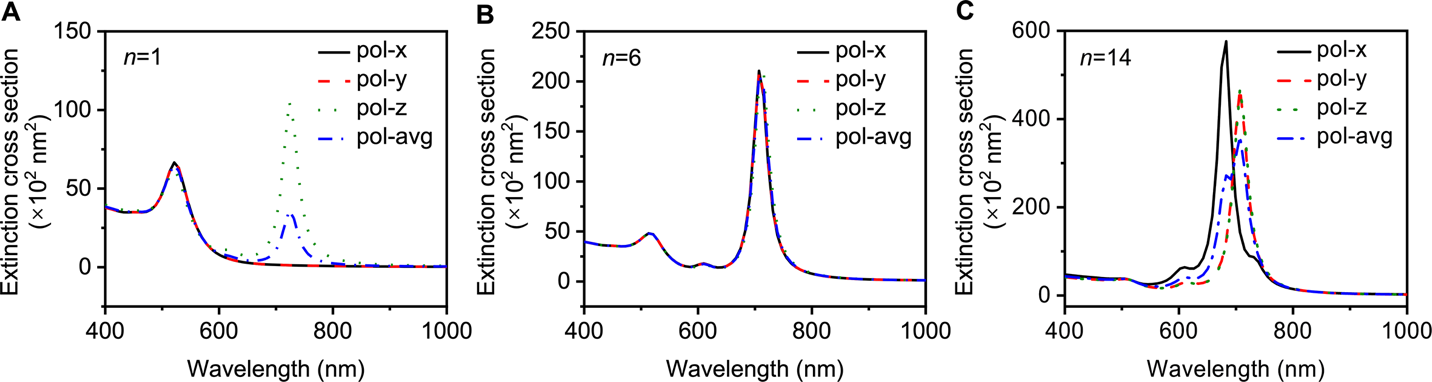

**Fig. S16 BEM simulation of the extinction cross-section for a single AuNU with various tip numbers.** (A) *n* = 1, (B) *n* = 6, and (C) *n* = 14. Note that the polarization directions are applied along x-, y-, and z- directions. The average extinction cross-section from three linear polarization directions is calculated for each case.

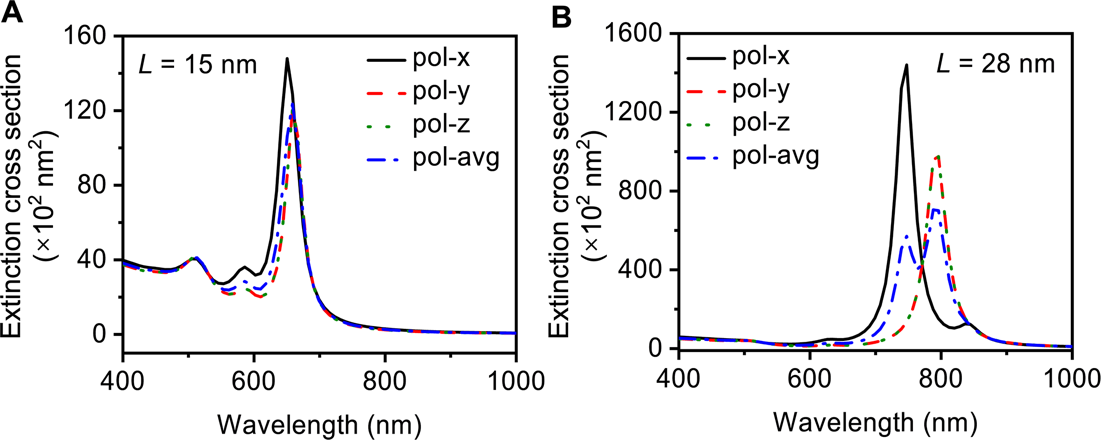

**Fig. S17 BEM simulation of the extinction cross-section for a single AuNU with various tip lengths.** (A) *L* = 15 nm, and (B) *L* = 28 nm. Note that the polarization directions are applied along x-, y-, and z- directions. The average extinction cross-section from three linear polarization directions is calculated for each case.

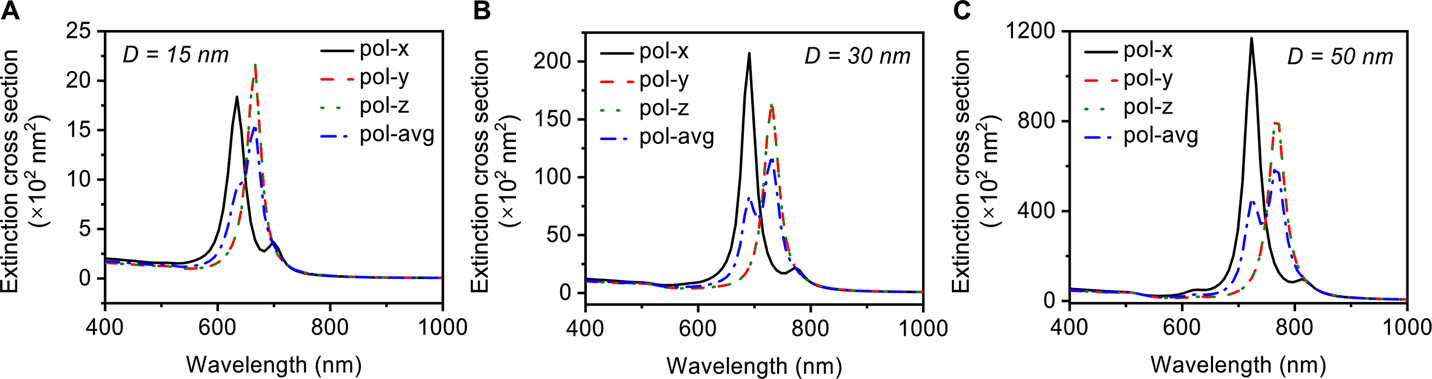

**Fig. S18 BEM simulation of the extinction cross-section for a single AuNU with different core sizes.** (A) *D* = 15 nm, (B) *D* = 30 nm, and (C) *D* = 50 nm. Note that the polarization directions are applied along x-, y-, and z- directions. The average extinction cross-section from three linear polarization directions is calculated for each case. The ratio of *D*/*L* was set as 0.5.

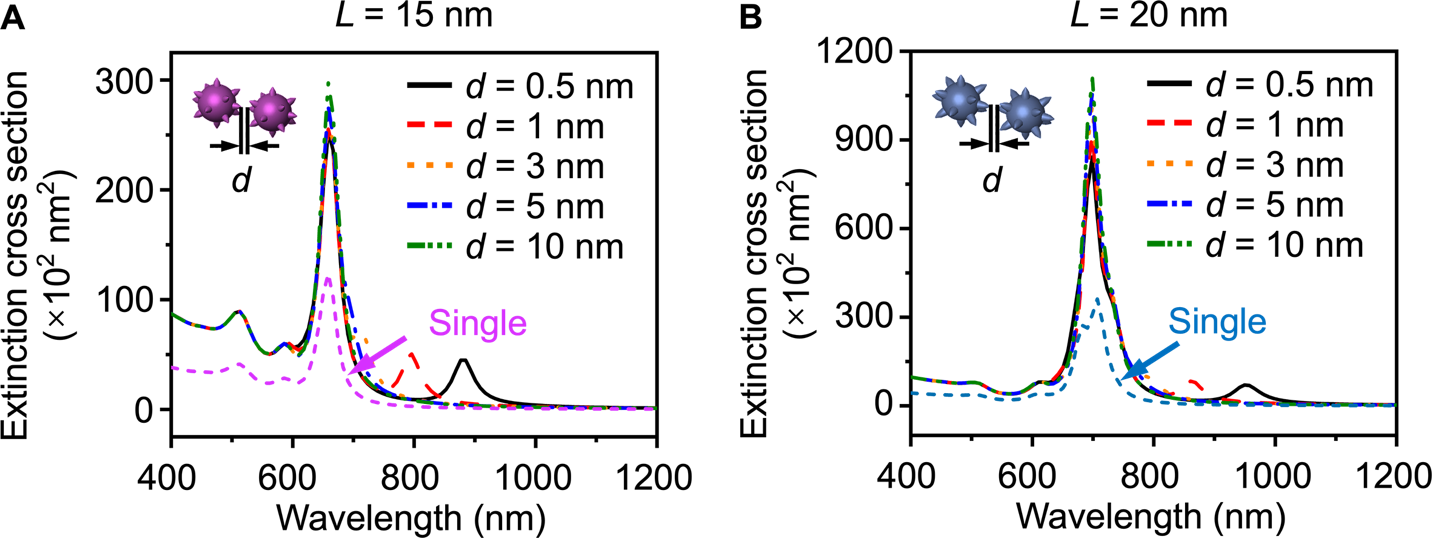

**Fig. S19 BEM simulation of the plasmonic coupling properties for coupled AuNUs.** Simulated extinction cross-section at different interparticle distance *d* for AuNUs with tip lengths of (A) *L* = 15 nm and (B) *L* = 20 nm. *d* is defined as tip-to-tip distance. The core size and tip number in the models are fixed with *D* = 50 nm and *n* = 14. Increasing the tip length leads to improved plasmonic coupling strength.

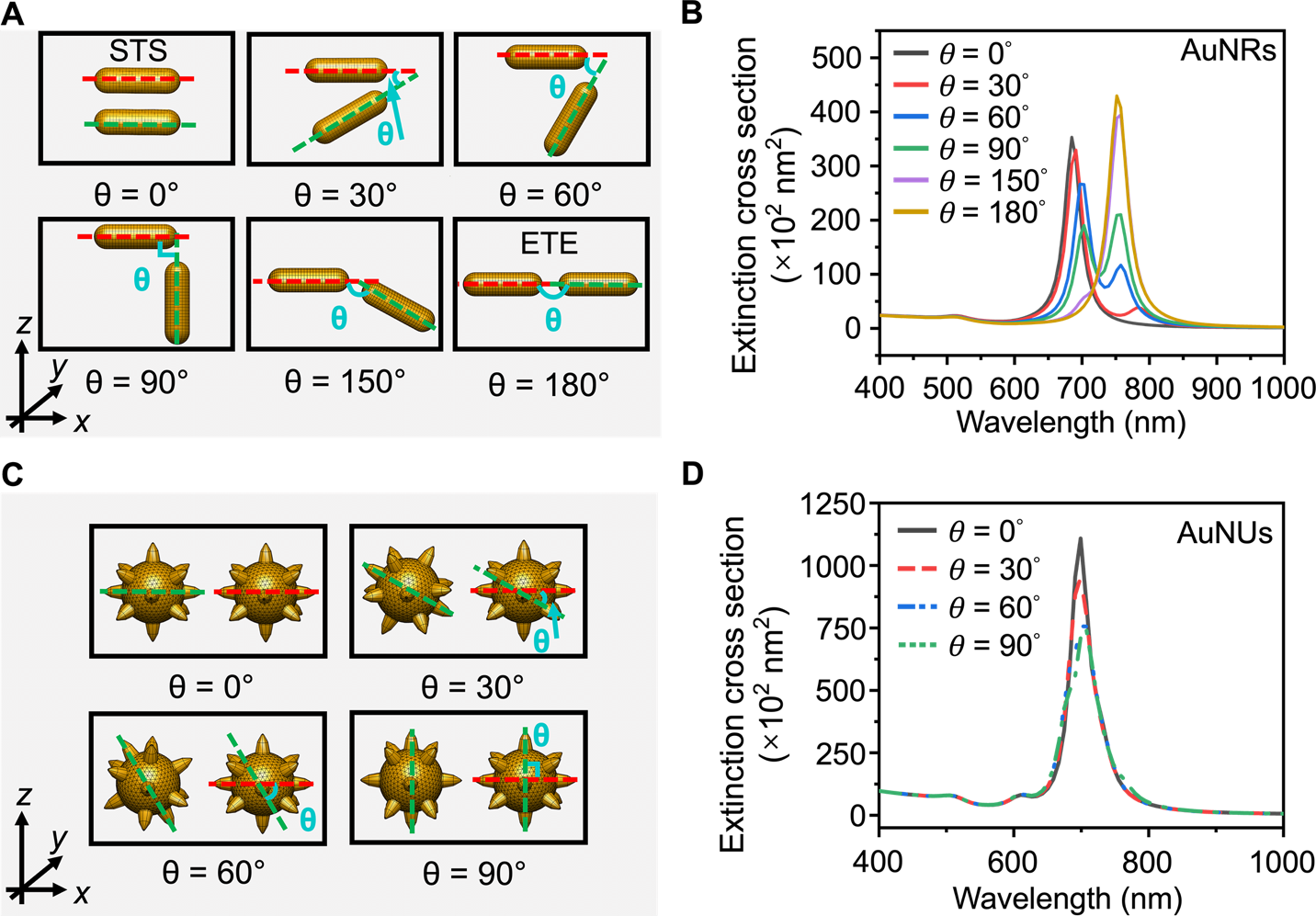

**Fig. S20 Influence of coupling angle between two anisotropic NPs on their optical properties.** Simulation models (A, C) and calculated extinction cross-section (B, D) for two coupled AuNRs and AuNUs at different coupling angles *θ*. The interparticle distance *d* was kept at 10 nm. Note the same results can be found for AuNUs with *θ* beyond 90° due to the symmetry.

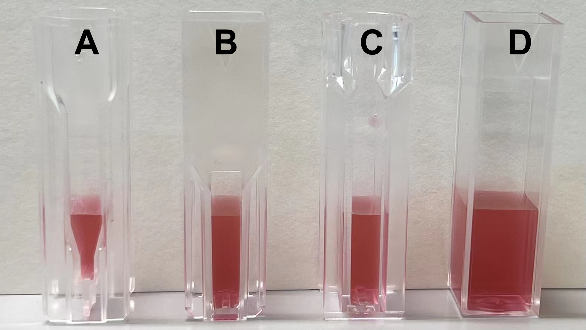

**Fig. S21 Disposable spectrometer cuvettes (Fisher Scientific) made of clear plastic for smartphone testing.** Measuring volumes (MV) are flexible. (A) MV = 300 µL, (B, C) MV = 600 µL, and (D) MV = 1.2 mL.
